# Comparison of various transcatheter aortic valves for aortic stenosis – a network meta-analysis of randomized controlled trials

**DOI:** 10.1101/2022.10.16.22281140

**Authors:** Emily Hiltner, Monarch Shah, Derek Schwabe-Warf, Bruce Haik, Abdul Hakeem, Mark Russo, Ankur Sethi

**Affiliations:** Robert Wood Johnson University Hospital, New Brunswick, NJ; St. Peter’s University Hospital, New Brunswick, NJ

**Keywords:** Transcatheter aortic valve replacement, aortic valve replacement, aortic valve disease, meta-analysis

## Abstract

**Objectives:** Our aim was to compare available transcatheter aortic valve replacement (TAVR) valves using direct and indirect evidence from randomized controlled trials (RCT).

**Background:** TAVR is now an established treatment for majority of patients with severe aortic stenosis. However, there is limited data comparing various valves.

**Methods:** We performed a systematic search of electronic databases for RCT comparing a TAVR valve to a valve or surgery. A Bayesian network meta-analysis was performed to compile evidence from both direct and indirect comparisons at 30 days and at one year.

**Results:** Twelve studies with 10,307 patients eligible for TAVR met the criteria and were included. Self-expanding valve CoreValve type (SEV_C) is associated with higher risk of pacemaker implantation and use of >1 valve, SEV Accurate type (SEV_A) is associated with higher risk of ≥ moderate aortic regurgitation (AR) and death, and mechanically expandable valve (MEV) is associated with lower risk of ≥ moderate AR but higher risk of pacemaker at 30 days, SEV_C and MEV were associated with higher pacemaker rates compared balloon expandable valve (BEV) at 1 year. There is no difference among the valves in stroke at 30 days and 1 year.

**Conclusions:** At 30 days, BEV was superior on one or more outcomes of mortality, pacemaker implantation, >1 valve implantation, and ≥ moderate AR compared to other valves except the higher rate ≥?moderate AR compared to MEV. At one year, BEV was associated with lower odds of pacemaker implantation compared to SEV_C and MEV but not different on other end points.

## Introduction

The last decade has seen unprecedented research and development in the transcatheter valve therapies for the treatment of severe aortic stenosis (AS). Multiple large, randomized trials have clearly established transcatheter aortic valve replacement (TAVR) as either preferred treatment or suitable alternative across the whole spectrum of patients with AS [1-3]. The most recent 2020 American Heart Association (AHA)/American College of Cardiology (ACC) guidelines have given TAVR a Class I recommendation for the treatment for symptomatic AS in patients older than 65 years, and suitable valve and vascular anatomy regardless of the surgical risk [4]. Even though there is widespread acceptance of TAVR, the choice of most suitable transcatheter valve for AS is far from clear. There are several different types of transcatheter valves in use or under investigation for severe AS in different parts of the world. At least three TAVR valves are approved for commercial use in the United States. There are several differences amongst the available TAVR valves including mode of deployment, intra-annular versus supra-annular placement, length of the valve frame, leaflet type and thickness which may impact both periprocedural and long-term outcomes. There are few randomized trials providing head-to-head comparisons amongst the various TAVR valves [5-9]. Therefore, we performed a network meta-analysis to compile both direct and indirect evidence comparing outcomes amongst TAVR valves for severe symptomatic native aortic valve stenosis.

## Materials and Methods

### Search strategy and inclusion criteria

We performed a systematic search of electronic databases, PubMed/Medline, Embase and Cochrane Central Register of Controlled Trials (CENTRAL), for randomized controlled trials comparing TAVR to control in patients with AS. The following key words in various combinations were used – “transcatheter aortic valve replacement”, “TAVR”, “transcatheter aortic valve implantation”, “TAVI”, “aortic stenosis”, “clinical trial”. In addition, we searched clinical trials.gov and acc.org for studies and follow up not yet published. The bibliographies of review articles and systematic reviews were hand searched for any relevant studies. Last search was performed on October 30, 2021. No language restriction was enforced. All human studies using a transcatheter aortic valve in a randomized fashion for treatment of AS were eligible. Non-randomized and observational studies were excluded. Studies using more than one type of transcatheter valve in a treatment arm were also excluded.

### Data extraction

Two investigators independently evaluated studies for inclusion in the meta-analysis. Studies meeting the inclusion criteria were independently reviewed and relevant study characteristics and end points were extracted and entered in preset form. Any discrepancies were resolved with consensus or help of a third author. We used the revised Cochrane risk of bias tool for randomized trial to assess the quality of included studies [10]. This meta-analysis was conducted in accordance with the Preferred reporting items for systemic reviews and meta-analysis (PRISMA) statement [11]. We used the tool developed by GRADE working group to rate the confidence in the estimates of network meta-analysis for all outcomes [12].

### End points

Death and stroke at 30 days and 1 year were primary clinical end points. Other valve related end points assessed were ≥ moderate aortic regurgitation (AR), pacemaker implantation, valve reintervention, and use of >1 transcatheter valve during the primary procedure.

### Statistical analysis

We used intention to treat analysis for all outcomes included in the meta-analysis. A random effect Bayesian network meta-analysis was performed with uninformative prior using 20,000 adaptive iteration and 100,000 simulations. Model convergence was evaluated using Brooks-Gelman-Rubin plot and potential scale reduction factor (shrink factor). A model with decreasing variation over increasing number of simulations and shrink factor < 1.05 was considered to have achieved convergence. We used nodesplit models to assess inconsistency between the direct and indirect evidence for each outcome. A p value < 0.05 was considered an evidence of inconsistency. To preserve transitivity assumption to the extent possible, we excluded patients undergoing transapical TAVR. Furthermore, to account for difference in baseline risk among the studies, we performed a network meta-regression of mean STS score over the odds of mortality after different TAVR valves at 30 days and 1 year. We assessed the heterogeneity among the included studies using global I^2^ statistic for each outcome. However, to be conservative and allow for differences between the studies, we used random effect model for our estimates. Odds ratio with 95% credible interval (95% CrI) was used as summary estimate. A 95% CrI which did not contain unity was considered statistically significant. The statistical analysis was conducted using the GeMTC package in R 3.1.2 statistical software using JAGS (Just another Gibbs Sampler) as the sampler [13].

## Results

Twelve studies [1-3,5-9,14-17] with a total of 10,307 patients met the inclusion criteria. The search strategy and selection process are shown in Figure 1. The following valves were compared: balloon expandable valve – SAPIEN /SAPIEN XT/SAPIEN 3 (Edwards Lifesciences, Irvine, CA, USA) (BEV); self-expanding valve – CoreValve/Evolut R/Evolut Pro (Medtronic Inc, Minneapolis, MN, USA) (SEV_C); self-expanding valve – Accurate Neo (Boston Scientific, Marlborough, MA, USA) (SEV_A), and mechanically expanding valve – LOTUS (Boston Scientific, Marlborough, MA, USA) (MEV). TAVR was performed via transfemoral access in all except a minority of patients (Table 1). The mean age of patients ranged from 73 to 84 years. The Society of thoracic Surgeons (STS) scores range from 1.9% to 11.8%. Other relevant study characteristics are shown in Table 1. The network plot of trials contributing direct and indirect evidence comparing different TAVR valves is shown in Figure 1S.

**Table 1.** Characteristics of included studies. Accurate Neo (Boston Scientific, Marlborough, MA, USA) Self-expanding valve Accurate neo type, CABG – Coronary artery bypass graft, CAD – Coronary artery disease, CoreValve/Evolut R/Evolut Pro (Medtronic Inc, Minneapolis, MN, USA) Self-expanding valve CoreValve type, LOTUS (Boston Scientific, Marlborough, MA, USA) Mechanically expanding valve, PAD – Peripheral artery disease, NR – Not reported, SAPIEN /SAPIEN XT/SAPIEN 3 (Edwards Lifesciences, Irvine, CA, USA) Balloon expandable valve, SAVR – Surgical aortic valve replacement, STS – Society of thoracic surgeons, TAVR – Transcatheter aortic valve replacement, TF – Transfemoral, Y – Years.

|  |  | INCLUSION<br>CRITERIA | PERIOD | RAL ACCESS<br>(%) | (IN YEARS) |  |  |  | (%) |  | (%) |  | (%) |  | FIBRILLATION<br>(%) |  | (%) |  |
| --- | --- | --- | --- | --- | --- | --- | --- | --- | --- | --- | --- | --- | --- | --- | --- | --- | --- | --- |
|  |  |  |  |  | Comparator<br>1 | Comparator<br>2 | Comparator<br>1 | Comparator<br>2 | Comparator<br>1 | Comparator<br>2 | Comparator<br>1 | Comparator<br>2 | Comparator<br>1 | Comparator<br>2 | Comparator<br>1 | Comparator<br>2 | Comparator<br>1 | Comparator<br>2 |
| SCOPE I (5) | SAPIEN 3 vs<br>ACURATE NEO | >75 Y patients<br>with increased<br>risk (STS score<br>> 10 or other<br>criteria) and<br>suitable for TF<br>access | 2017-19 | 100 | 83 | 82.6 | 3.4 | 3.7 | 60 | 59 | 8 | 8 | 32 | 29 | 37 | 36 | 11 | 12 |
| CHOICE (6) | SAPEIN XT vs<br>COREVALVE | High risk<br>patients (STS<br>score ≥ 10 or<br>other criteria)<br>and suitable<br>for TF access | 2012-13 | 100 | 81.9 | 79.6 | 5.6 | 6.2 | 60.3 | 65.8 | 15.7 | 12.5 | 31.4 | 26.7 | 33.3 | 24.8 | 16.5 | 18.3 |
| SOLVE TAVI (7) | SAPIEN 3 vs<br>CoreValve<br>(EVOLUT R) | >75 Y patients'<br>high risk for<br>SAVR suitable<br>for TF access | 2016-18 | 100 | 81.5 | 81.7 | 4.7 | 4.9 | 52.7 | 58 | 8.2 | 11.9 | 31.1 | 36.2 | 42.5 | 47 | 11.8 | 13.3 |
| SCOPE II (8) | CoreValve<br>(Evolut R/Pro)<br>vs ACURATE<br>neo | >75 Y TF TAVR<br>patient at<br>increased risk<br>for SAVR | 2017-19 | 100 | 82.9 | 83.4 | 4.5 | 4.6 | 38 | 43 | 4 | 7 | 29 | 27 | 32 | 35 | 10 | 8 |
| REPRISE III (9) | CoreValve<br>(Classic/Evolut<br>R) vs LOTUS | STS score ≥ 8<br>or other<br>criteria for<br>Increased or<br>extreme risk<br>for SAVR | 2014-15 | 100 | 82.9 | 82.8 | 6.9 | 6.7 | 73.4 | 71.5 | 20 | 16 | 32.6 | 30.9 | 31.6 | 35.1 | 25.7 | 31.1 |
| Evolut low risk<br>trial (14) | CoreValve<br>(Classic/Evolut<br>R/Pro) vs<br>SAVR | Predicted risk<br>of death after<br>SAVR < 3% | 2016-18 | 99 | 74 | 73.8 | 1.9 | 1.9 | NR | NR | 2.5 | 2.3 | 31.1 | 30.5 | 15.5 | 14.9 | 7.6 | 8.5 |
| NOTION (15) | CoreValve vs<br>SAVR | ≥ 70 Y<br>regardless of<br>risk | 2009-13 | 96.5 | 79.2 | 79 | 2.9 | 3.1 | NR | NR | NR | NR | 17.9 | 20.7 | 27.8 | 25.6 | 4.1 | 6.7 |
| SURTAVI (16) | CoreValve vs<br>SAVR | Intermediate<br>surgical risk<br>based on STS | 2012-16 | 93.6 | 79.9 | 79.7 | 4.4 | 4.5 | 62.6 | 64.2 | 16 | 17.2 | 34.1 | 34.8 | 28.1 | 26.5 | 30.8 | 29.9 |

Table 1 – Characteristics of included studies.
|  |  | score ≥ 3 but <15, and other factors |  |  |  |  |  |  |  |  |  |  |  |  |  |  |  |  |
| --- | --- | --- | --- | --- | --- | --- | --- | --- | --- | --- | --- | --- | --- | --- | --- | --- | --- | --- |
| PARTNER 3 (3) | Sapien 3 vs SAVR | Low risk for SAVR (STS < 4) suitable for TF access | 2016-17 | 100 | 73.3 | 73.6 | 1.9 | 1.9 | 27.7 | 28 | nr | nr | 31.2 | 30.2 | 15.7 | 18.8 | 6.9 | 7.3 |
| PARTNER 2 (TF cohort) (2) | SAPIEN XT vs SAVR | Intermediate surgical risk based on STS score ≥ 4 but <8, or other factors | 2011-13 | 100 | 81.5 | 81.7 | 5.8 | 5.8 | 69.2 | 66.5 | nr | nr | 37.7 | 34.2 | 31 | 35.2 | 27.9 | 32.9 |
| PARTNER 1A (TF cohort) (1) | SAPIEN vs SAVR | High surgical risk (STS score > 10) and at least 15% risk of death by 30 days | 2007-09 | 100 | 83.6 | 84.5 | 11.8 | 11.7 | 74.9 | 76.9 | 42.6 | 44.2 | nr | nr | 40.8 | 42.7 | 43 | 41.6 |
| US CoreValve high risk trial (17) | CoreValve vs SAVR | Predicted risk of operative mortality after SAVR ≥15% but < 50% | 2011-12 | 84 | 83.2 | 83.5 | 7.3 | 7.5 | 75.4 | 76.3 | 29.7 | 30.2 | 34.5 | 42.9 | 41 | 47.5 | 41.7 | 42.5 |

**Figure 1.**
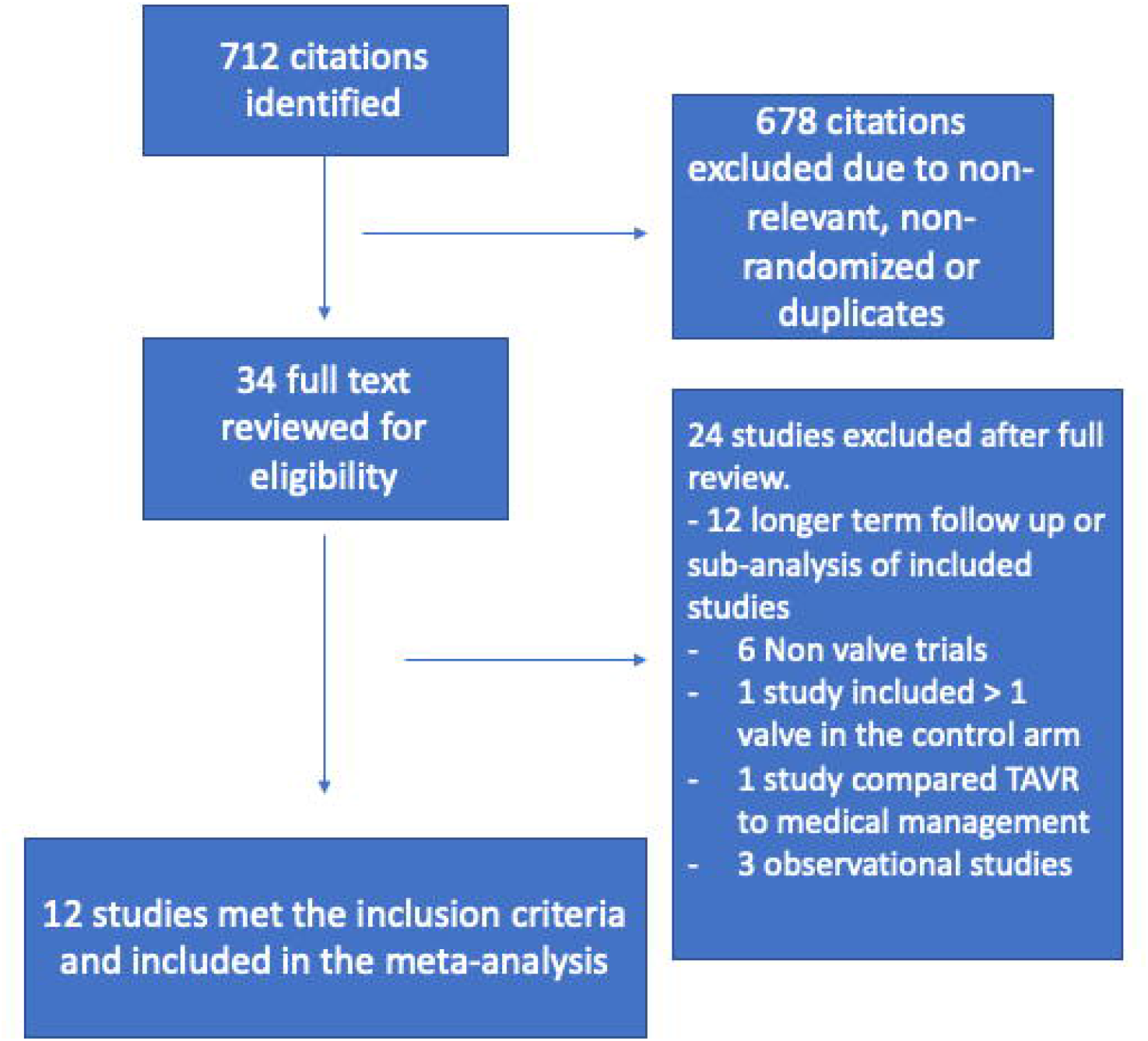
Flow diagram depicting search strategy for the network meta-analysis.

At 30 days, SEV_A was associated with higher 30-day mortality compared to BEV (OR = 3.0 95% CrI 1.1-8.7) as shown in Figure 2A, however there was no difference in mortality at 1 year in other valves compared to BEV (Figure 2B). Furthermore, the network meta-regression did not show any effect of baseline STS score on odds ratio after SEV_C, SEV_A and MEV compared to BEV at 30 days and 1 year follow up (Figure 2S and 3S). There was no difference in stroke at 30 days and 1 year after SEV_C, SEV_A and MEV compared to BEV as shown in Figure 3A and 3B. The models for mortality and stroke showed appropriate convergence and no evidence of inconsistency was found on node splitting models except for stroke at 1 year where there is inconsistency for the comparison SEV_C and BEV (Figure 4S-11S).

**Figure 2.**
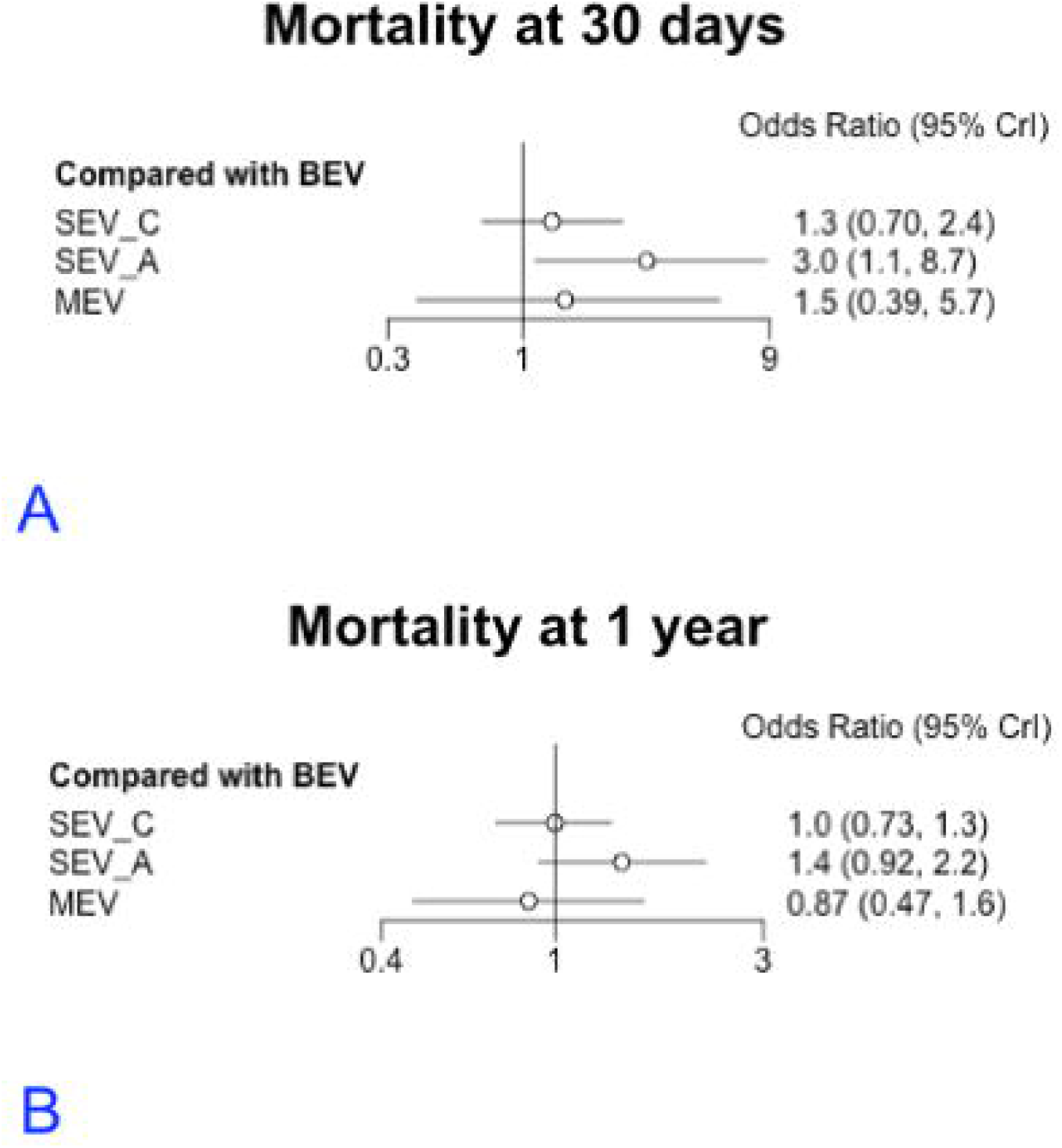
Forest plot showing of odds ratio of mortality after different valves compared to balloon expandable valve (BEV) A) at 30 days and B) at 1 year. MEV – Mechanically expanding valve, SEV_A – Self expanding valve Accurate Neo type, SEV_C – Self expanding valve CoreValve type.

**Figure 3.**
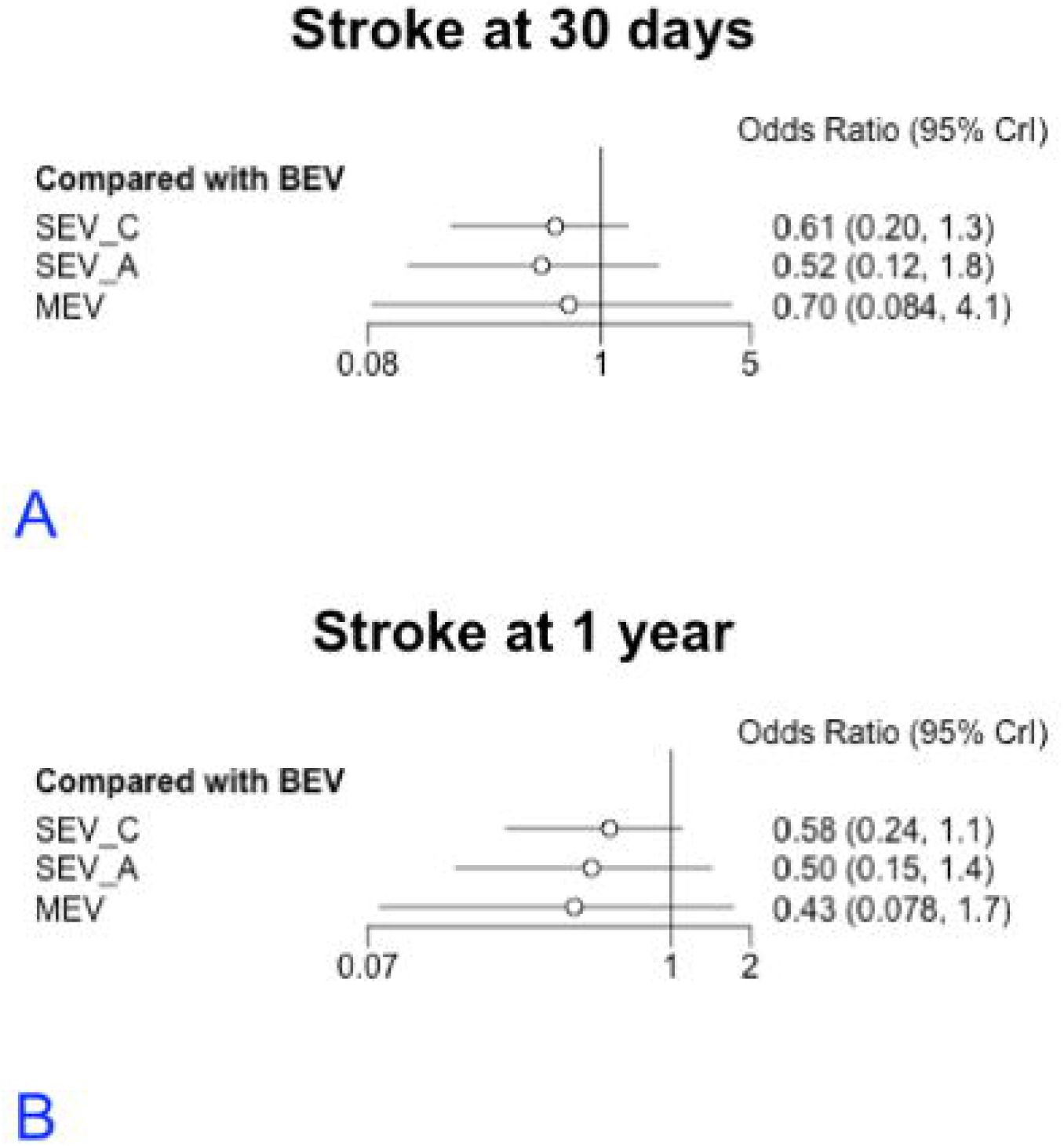
Forest plot showing of odds ratio of stroke after different valves compared to balloon expandable valve (BEV) A) at 30 days and B) at 1 year. MEV – Mechanically expanding valve, SEV_A – Self expanding valve Accurate Neo type, SEV_C – Self expanding valve CoreValve type.

**Figure 4.**
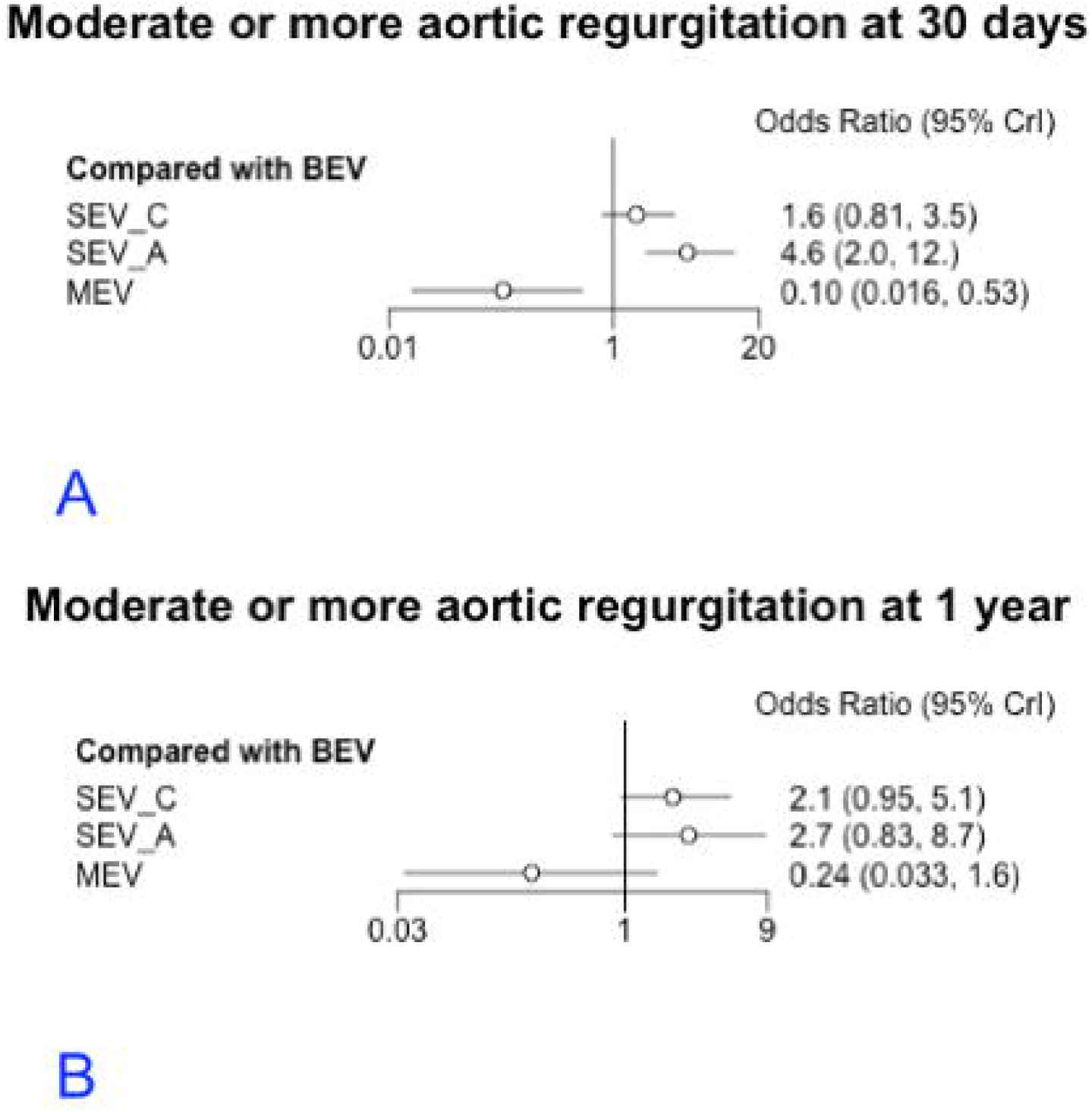
Forest plot showing of odds ratio of ≥ moderate aortic regurgitation after different valves compared to balloon expandable valve (BEV) A) at 30 days and B) at 1 year. MEV – Mechanically expanding valve, SEV_A – Self expanding valve Accurate Neo type, SEV_C – Self expanding valve CoreValve type.

At 30 days follow up, moderate or severe AR was significantly more common after SEV_A and less common after MEV compared to BEV (Figure 4A). There was no difference in 30 days ≥ moderate AR between SEV_C and BEV. There was no difference in ≥ moderate AR between the other valves and BEV at one year (Figure 4B). The models showed appropriate convergence and no evidence of inconsistency was noted in node splitting models (Figure 12S-15S). Also, there was no difference in ≥ moderate paravalvular regurgitation after SEV_C (OR = 2.3 95% CrI 0.98-6.5), SEV_A (OR = 2.8 95% CrI 0.80-11) and MEV (OR = 0.26 95% CrI 0.03-2.3) compared BEV at one year (Figure 16S). All studies except PARTNER 1B trial [1] used Valve Academic Research Consortium (VARC) consensus statement version 1 [14-15] or version 2 definitions for aortic regurgitation [18-19]. However, exclusion of PARTNER 1B trial did not significant effect on network estimate for any valve compared to BEV at 30 days (SEV_C OR = 1.8 95% CrI 0.87-4.3; SEV_A OR = 4.9 95% CrI 2-13; MEV OR = 0.12 95% CrI 0.018-0.67) and 1 year (SEV_C OR = 2.1 95% CrI 0.85-5.9; SEV_A OR = 2.6 95% CrI 0.76-10; MEV OR = 0.23 95% CrI 0.028-2) for ≥ moderate regurgitation.

Pacemaker implantation was significantly higher after SEV_C and MEV at 30 days (OR = 2.5 95% CrI 1.3-5.3; OR = 5.7 95% CrI 1.3-29) and one year (OR = 2.3 95% CrI 1.1-4.9; OR = 5.4 95% CrI 1.1-28) compared to BEV (Figure 5A and 5B). There was no difference in pacemaker implantation after SEV_A and BEV. The models showed appropriate convergence and no evidence of inconsistency based on node splitting model (Figure 17S – 20S).

**Figure 5.**
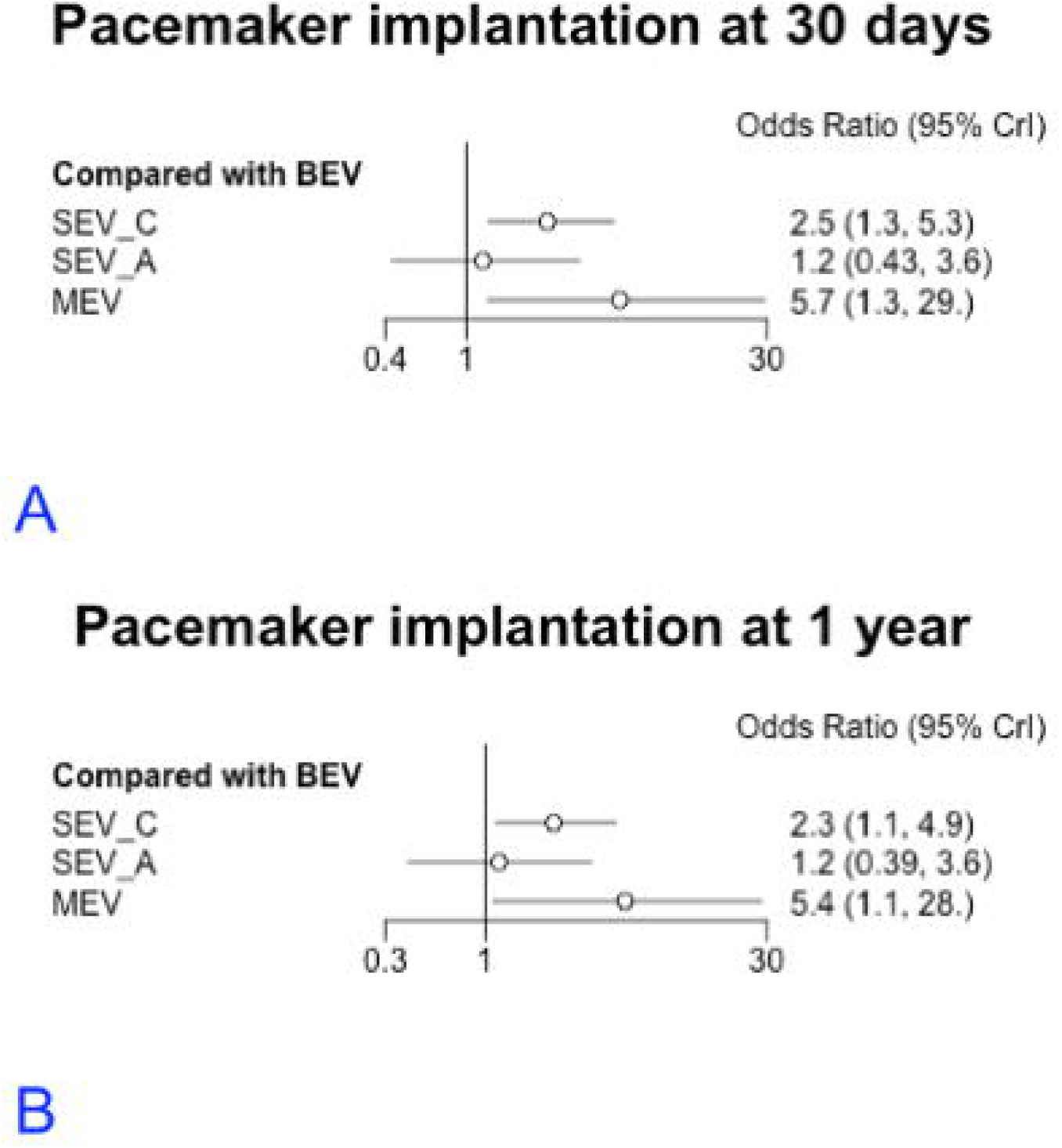
Forest plot showing of odds ratio of permanent pacemaker implantation after different valves compared to balloon expandable valve (BEV) A) at 30 days and B) at 1 year. MEV – Mechanically expanding valve, SEV_A – Self expanding valve Accurate Neo type, SEV_C – Self expanding valve CoreValve type.

The use of >1 transcatheter valve during the index procedure was significantly more common after SEV_C (OR = 16 95% CrI 1.4-3e^2^) but no different after SEV_A and MEV compared to BEV (Figure 21S). However, the potential scale reduction factor for this model did not reach the prespecified value of < 1.05 for the comparison between SEV_C and MEV (Figure 22S). After excluding the study using MEV, the model achieved convergence (Figure 23S) and continued to show that SEV_C was associated with > 1 valve use compared to BEV (Figure 6). There was no evidence of inconsistency as shown by node splitting model (Figure 24S).

**Figure 6.**
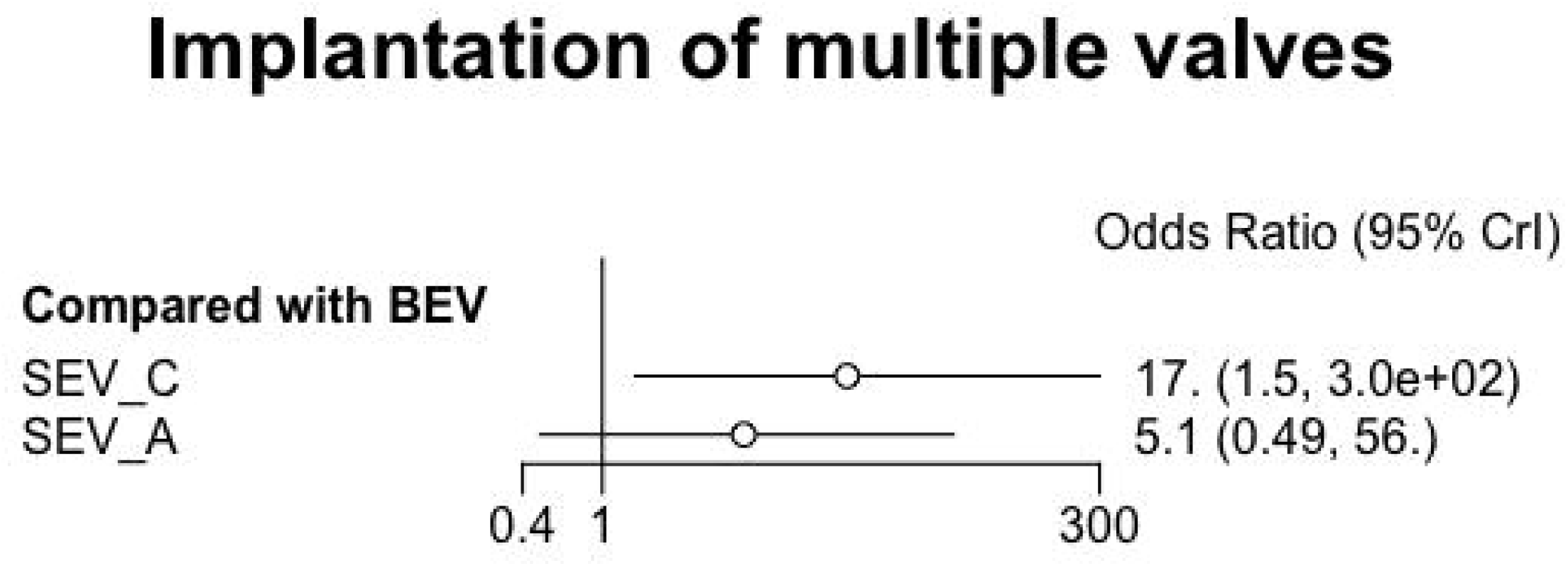
Forest plot showing of odds ratio use > 1 valve after different valves compared to balloon expandable valve (BEV). SEV_A – Self expanding valve Accurate Neo type, SEV_C – Self expanding valve CoreValve type.

The risk of bias evaluation for included studies is shown in Table 1S (supplement). The overall risk of bias was considered low for all included studies. Also, there was no significant heterogeneity based on global I^2^ statistic as shown in Table 2S (supplement). Based on the GRADE group recommendations there was high confidence in the estimates for mortality and pacemaker implantation at 30 days and 1 year for the comparison SEV_C versus BEV. All other estimates have moderate or low confidence mostly secondary to imprecision as shown in Table 3S (supplement).

## Discussion

We conducted a comprehensive network meta-analysis of randomized controlled trials comparing different TAVR valves used in the treatment of severe symptomatic AS. We found SEV_A had higher mortality and ≥ moderate AR compared to BEV at 30 days, but not at one year follow up. SEV_C and MEV were associated with significantly increased risk of pacemaker implantation at 30 days and one year compared to BEV. SEV_C was associated with higher risk of use >1 valve compared to BEV. Our study represents one of the largest meta-analyses comparing contemporary TAVR prostheses and reports both short and mid-term outcomes.

We found a significantly increased risk of mortality at 30-days with SEV_A compared to BEV; however, this increased risk was no longer present at 1 year follow up. This lack of significant difference at 1 year could be secondary to imprecision in our estimate rather than absence of effect. This finding is very intriguing and needs to be addressed by an adequately powered clinical trial. Furthermore, there was no significant difference in mortality between BEV and SEV_C, the two of the most used TAVR valves. Our results are consistent with prior meta-analyses and a large propensity matched analysis showing no difference in 30 days and 1 year mortality between BEV and SEV [21-23]. To account for baseline difference in surgical risk, we performed a network meta-regression which did not show any effect of baseline STS score on odds of mortality for all prosthesis types compared to BEV.

Moderate or greater paravalvular leak/regurgitation (PVL) after TAVR has been previously shown to be associated with higher risk of heart failure hospitalization and long-term mortality [2,25-27]. PVL is likely the most common mechanism of regurgitation within the 30 days of the procedure as demonstrated by very low incidence of ≥ moderate AR in the surgical arms of the included trials. However, it may not always be feasible to differentiate between the intra-valvular and paravalvular regurgitation based on transthoracic studies performed at follow up in the included studies. We found at 30 days, there was increased risk of ≥ moderate AR in patients who received SEV_A and reduced risk in patients who received MEV compared to BEV. However, at 1 year follow up, there was no significant difference in the occurrence of ≥ moderate AR in all valves compared to BEV. Additionally, there was no difference in PVL between other valve types and BEV at 1 year follow up. A previous network meta-analysis of ten studies reported higher incidence of ≥ moderate PVL after SEV compared to BEV at 30 days [22]. However, this meta-analysis incorporated SEV_C and SEV_A in one group and did not include SCOPE (Safety and Efficacy Comparison of Two TAVI Systems in a Prospective Randomized Evaluation) II trial which showed significantly higher ≥ moderate AR in SEV_A compared to SEV_C. We did not find a significant difference in ≥ moderate AR between SEV_C and BEV, however, there was a strong trend of towards higher AR after SEV_C (Figure 3A and B). The CHOICE [6] and SOLVE TAVI [7] trials directly compared SEV_C to BEV, and the former found higher incidence of ≥ moderate AR whereas the later reported numerically higher ≥ moderate AR after SEV_C compared to BEV. Furthermore, FRANCE 2, a large national TAVR registry, reported SEV to be associated with higher risk ≥ moderate AR compared to BEV [26]. Therefore, additional studies with adequate power are required to resolve this issue particularly for the comparison of BEV to SEV_C and SEV_A. Also, despite lower risk of PVL compared to BEV and SEV, MEV is no longer available for use in the United States [27].

Our study found no significant difference in the risk of stroke with all valves studied compared to BEV. However, there was inconsistency between direct and indirect evidence particularly at one year. This inconsistency is likely due to the SOLVE TAVI trial, which directly compared SEV with BEV, and met the trial criteria for equivalence for the stroke reporting an incidence of 1% versus 6.9% respectively [7]. Whereas the incidence of stroke at one year after TAVR in the PARTNER 2 [2] and PARTNER 3 [3] trials, which compared BEV with SAVR, and SURTAVI trial [16] and the EVOLUT low risk trial [14], which compared SEV with SAVR, were 8%, 1.2%, 5.5% and 4.1% respectively. Furthermore, in a propensity matched analysis of >12,000 patients from 10 different studies (The CENTER-collaboration), BEV was associated with lower risk of stroke compared to SEV_C [23]. Therefore, we believe the equipoise between the valves in terms of stroke continues.

Permanent pacemaker (PPM) placement after TAVR has been associated with increased heart failure admissions, left ventricle systolic dysfunction and, in some studies, higher mortality [23-25]. As indications for TAVR extend to lower risk patient populations, it has become increasingly important to avoid this post-procedural complication. We found significantly increased risk of PPM placement in patients who received SEV_C and MEV compared to BEV. The previous network meta-analysis similarly reported increased risk of PPM placement with SEV compared to BEV [22]. Furthermore, the CENTER-collaboration found that SEV_C is associated with three-fold higher risk of PPM compared to BEV [23]. The continuous radial force of the nitinol frame in the SEV_C and the potential for deeper implantation leading to persistent AV node and bundle mechanical compression and injury have been postulated to explain these between-valve differences in PPM rates [31-32].

As per VARC consensus statement, successful delivery of a single functioning TAVR valve in a correct anatomical position without any major complication is required for technical success [32]. We found that use >1 valve is more common after SEV_C compared to BEV. Similarly in the FRANCE 2 registry, the use of > 1 valve occurred in 3.5% patients undergoing SEV_C compared 1.4% patients undergoing BEV [33]. The differences in deployment process, metallic frame, anchoring mechanism and delivery system may possibly explain the lower technical success of SEV_C compared to BEV [33].

Several limitations of our study need to be mentioned. The compilation of direct and indirect evidence in network meta-analysis is based on transitivity assumption. We systematically tested and reported inconsistency, if any, for each reported outcome using node-splitting model. We purposefully excluded the trans-apical cohort of prior BEV studies to preserve transitivity. Furthermore, to assess the effect of differences in baseline risk we performed a network meta-regression of mean STS score on odds ratio for mortality. In addition to allow for heterogeneity among the studies we performed a random effect meta-analysis which usually leads to wider confidence intervals, therefore, may not detect small differences in the effect. Because of the limited number of studies and use of more than one iteration of valves we were not able to compare just the latest iteration of BEV to SEV_C.

## Conclusion

At 30 days, BEV was superior on one or more outcomes of mortality, pacemaker implantation, >1 valve implantation, and ≥ moderate AR compared to other valves except the higher rate ≥ moderate AR compared to MEV. There is no significant different between the available valves in terms of mortality, stroke, ≥moderate regurgitation at 1 year, however, BEV is associated with lower risk of PPM compared to SEV_C and MEV.

## Data Availability

All data produced in the present work are contained in the manuscript.

## Supplementary Table Legends

Table 1S – Risk of bias evaluation of the included studies using Cochrane risk of bias tool.

Table 2S - Global I^2^ statistics to assess heterogeneity for all outcomes.

Table 3S – Grading the certainty of network estimates using GRADE group recommendations for network meta-analysis.

