## Supplemental Figures for "Comparison of various transcatheter aortic valves for aortic stenosis – a network meta-analysis of randomized controlled trials"

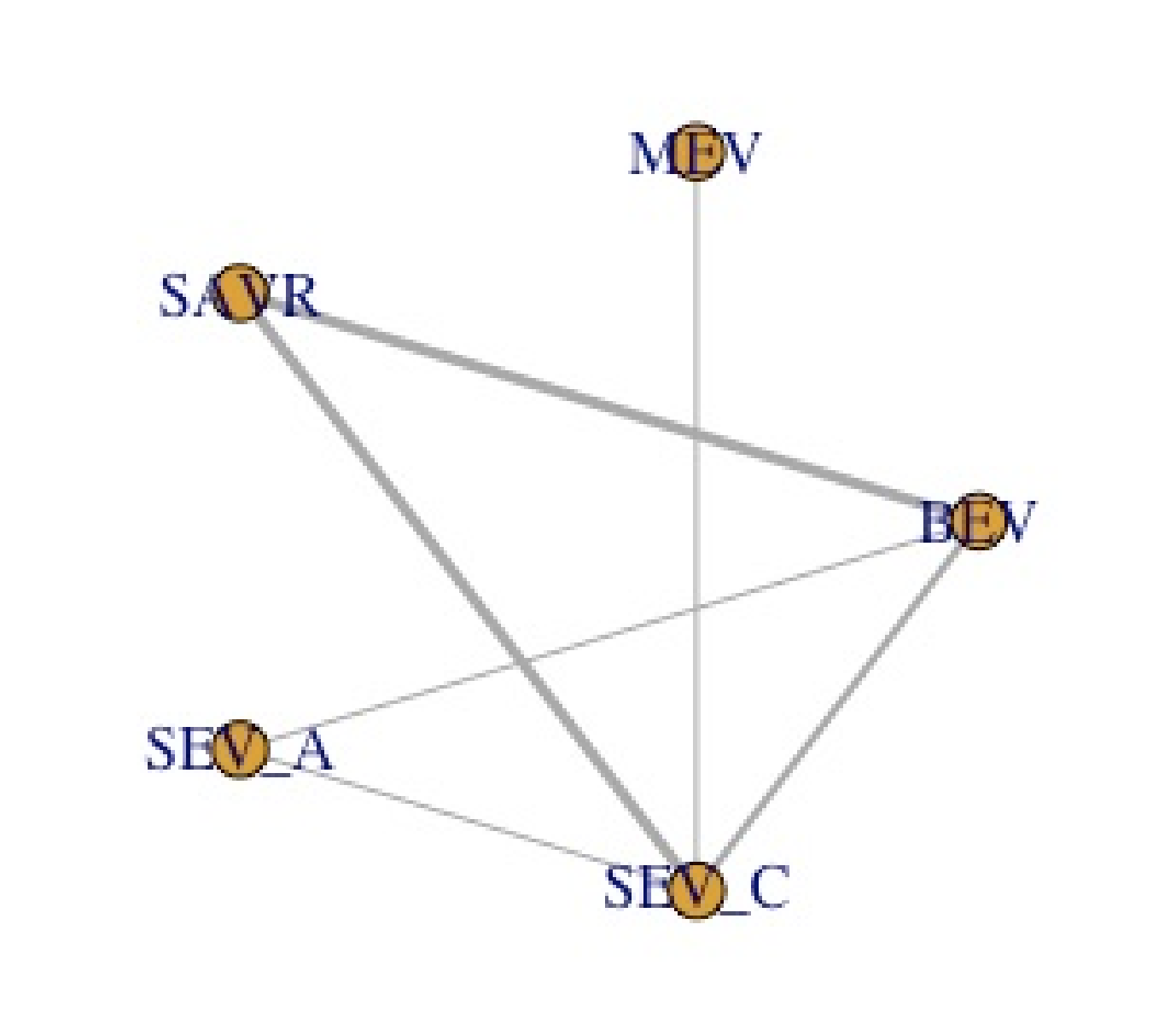

Figure 1S – Network plot depicting direct and indirect comparison of different valves. BEV – Balloon expandable valve, MEV – Mechanical expanding valve, SAVR – Surgical aortic valve replacement, SEV_A – Self expanding valve Accurate type, SEV_C -Self expanding valve CoreValve type

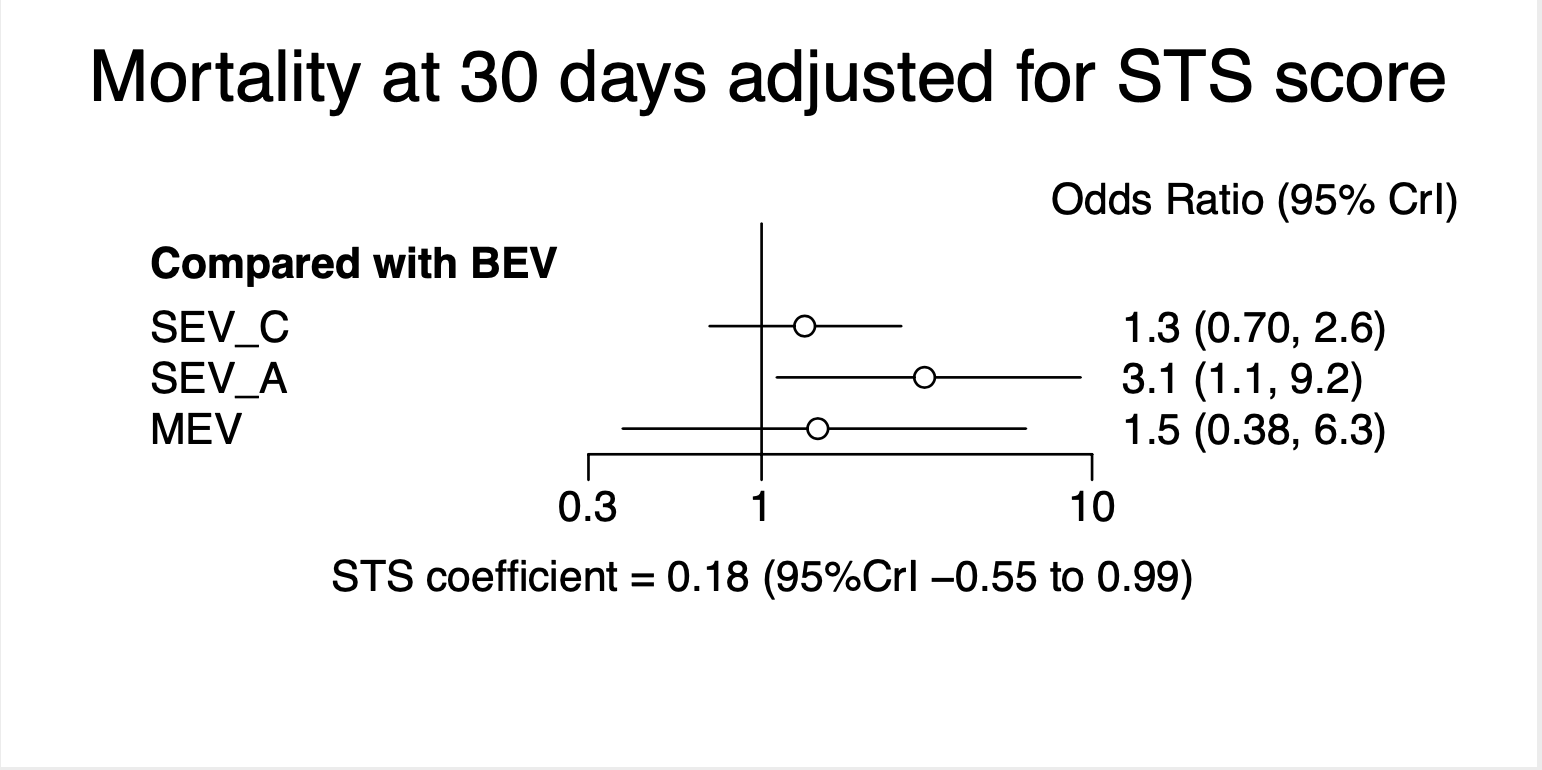

Figure 2S – Forest plot showing odds ratio of mortality at 30 days after SEV_C, SEV_A and MEV compared BEV adjusted for mean STS (Society of Thoracic Surgeons) score. Abbreviations same as figure 1S.

­
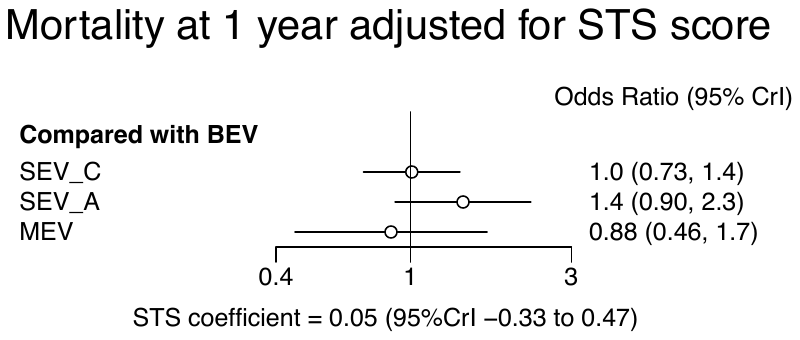

Figure 3S – Forest plot showing odds ratio of mortality at 1 year after SEV_C, SEV_A and MEV compared BEV adjusted for mean STS (Society of Thoracic Surgeons) score. Abbreviations same as figure 1S.

### Convergence diagnostics for mortality at 30 days

Potential scale reduction factors:

Point est. Upper C.I.

d.SEV_C.BEV	1	1

d.SEV_C.MEV	1	1

d.SEV_C.SAVR	1	1

d.SEV_C.SEV_A	1	1

sd.d	1	1

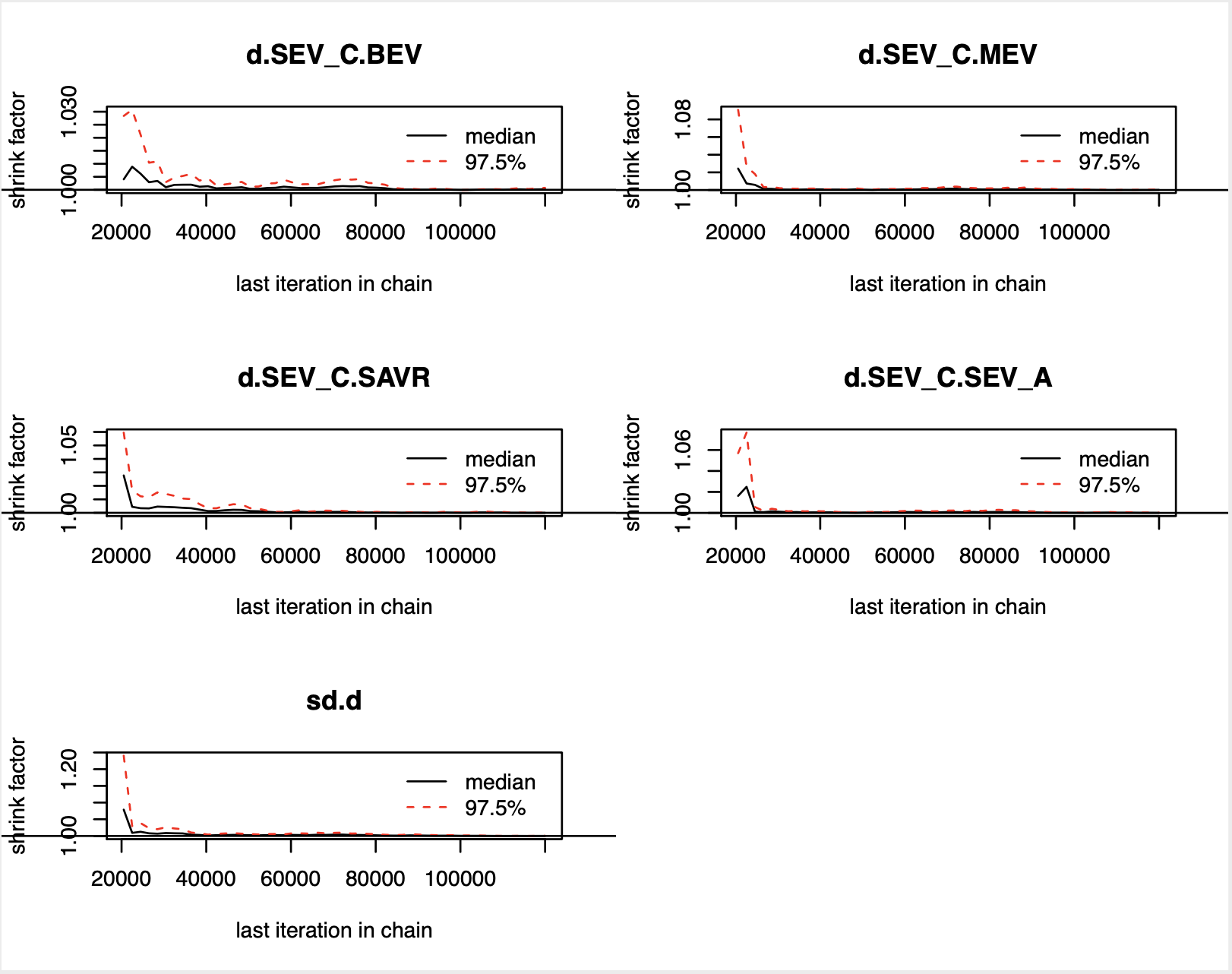

Figure 4S – Gelman-brooks-rubin convergence plot for 30 days mortality. Abbreviations same as figure 1S.

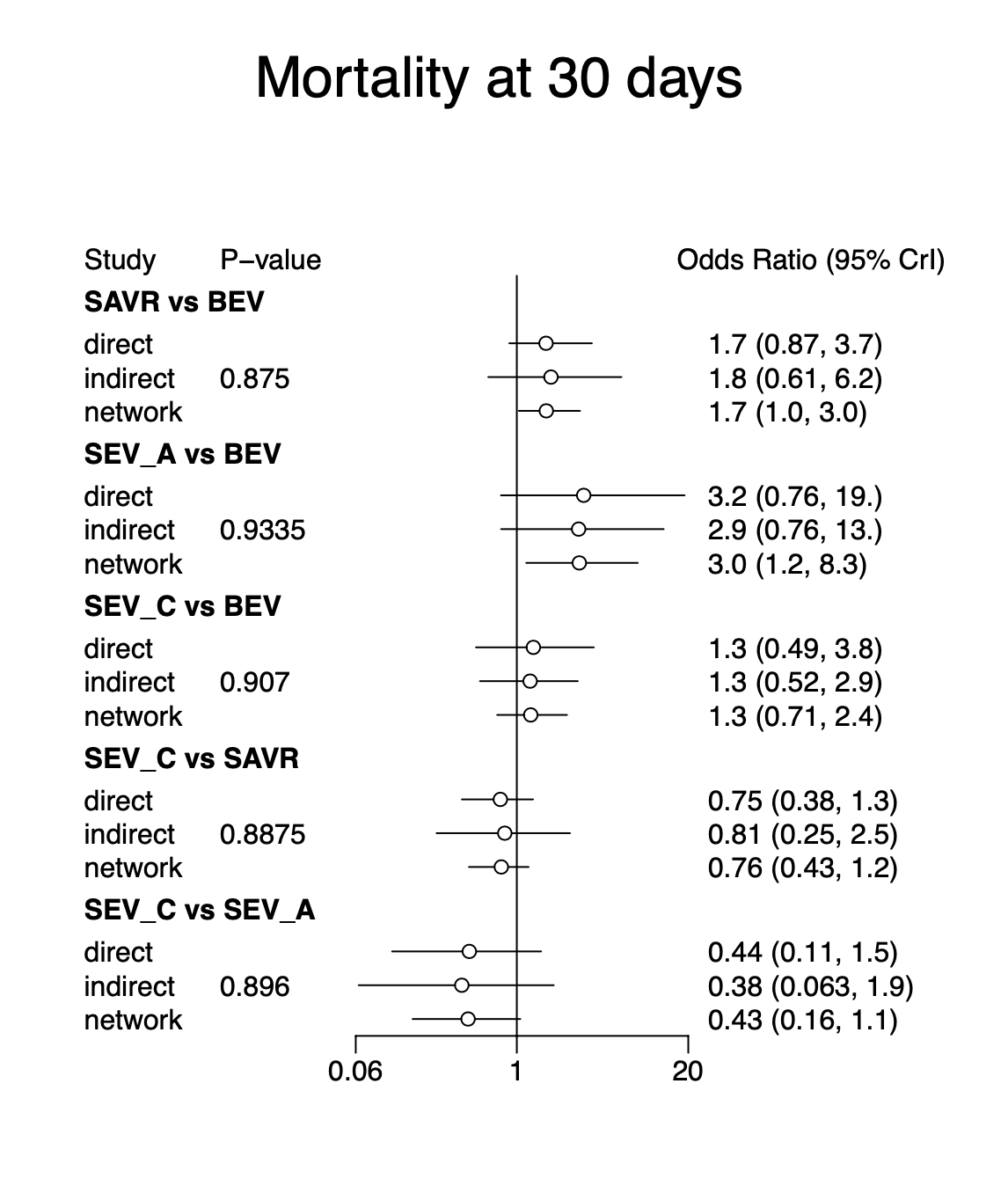

Figure 5S– Node splitting model for 30 days mortality after various TAVR valves. Abbreviations same as figure 1S.

### Convergence diagnostics for mortality at 1 year

Potential scale reduction factors:

Point est. Upper C.I.

d.SEV_C.BEV	1	1

d.SEV_C.MEV	1	1

d.SEV_C.SAVR	1	1

d.SEV_C.SEV_A	1	1

sd.d	1	1

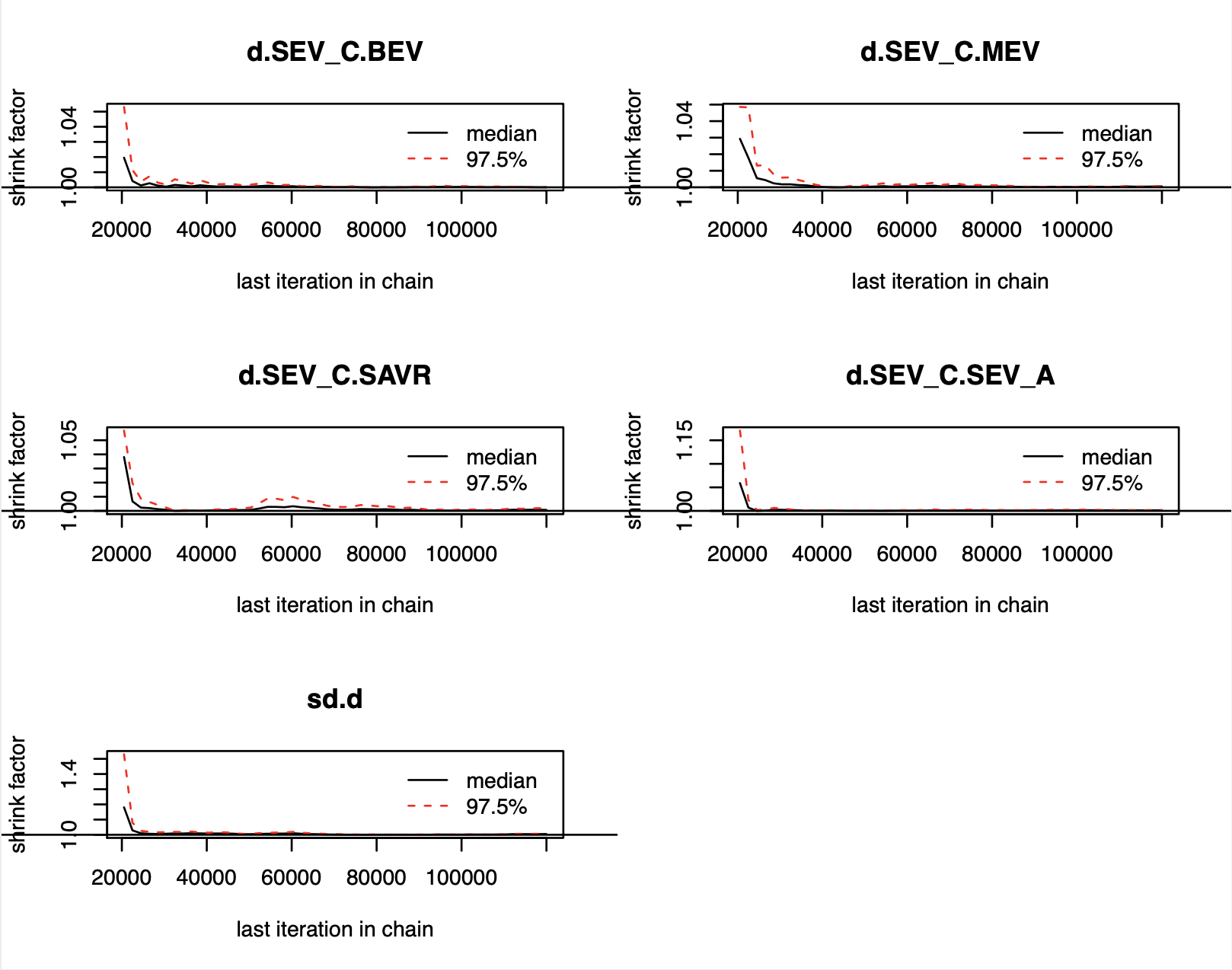

Figure 6S – Gelman-brooks-rubin convergence plot for one year mortality. Abbreviations same as figure 1S.

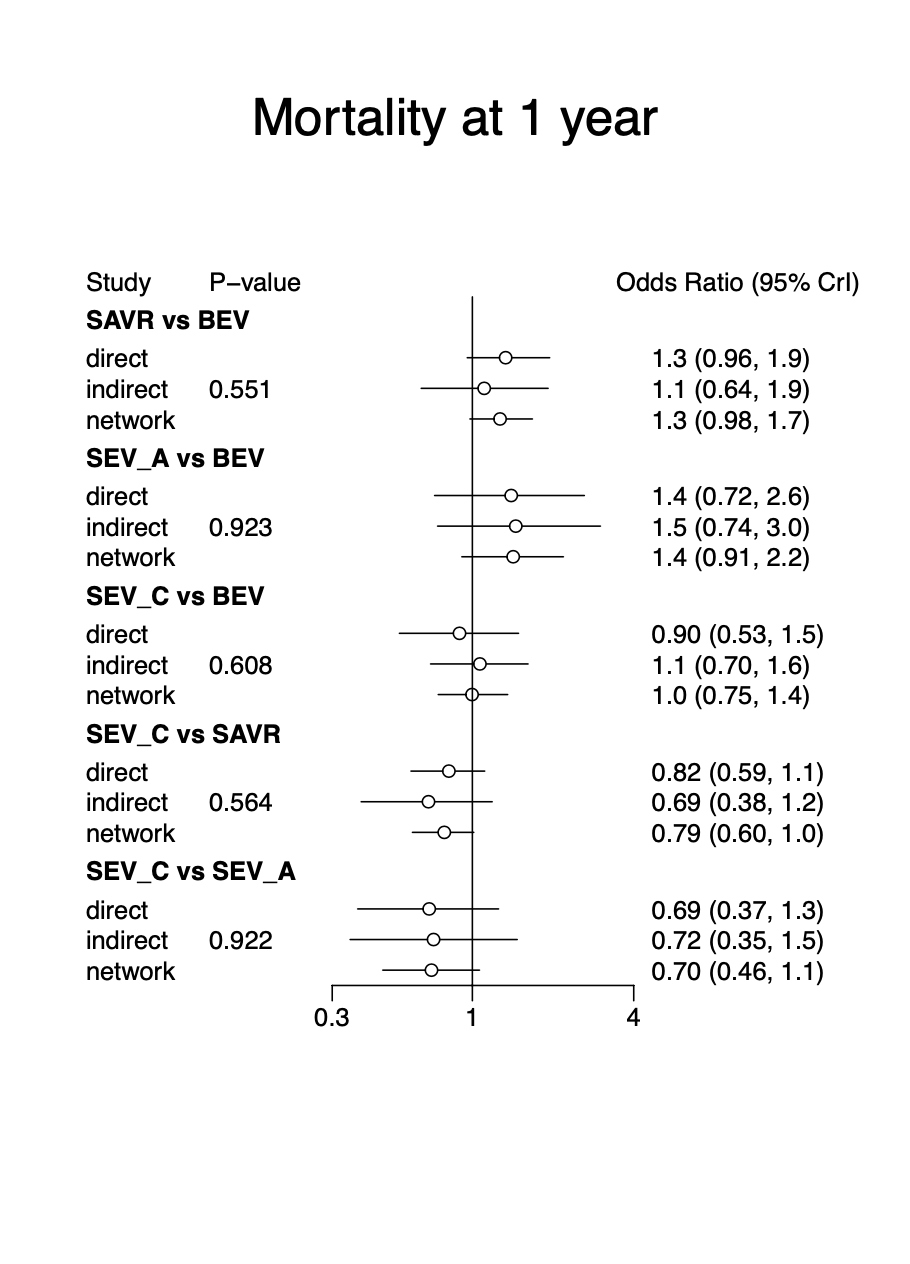

Figure 7S– Node splitting model for one year mortality after various TAVR valves. Abbreviations same as figure 1S.

### Gelman diagnostics for stroke at 30 days

Potential scale reduction factors:

Point est. Upper C.I.

d.SEV_C.BEV	1	1

d.SEV_C.MEV	1	1

d.SEV_C.SAVR	1	1

d.SEV_C.SEV_A	1	1

sd.d	1	1

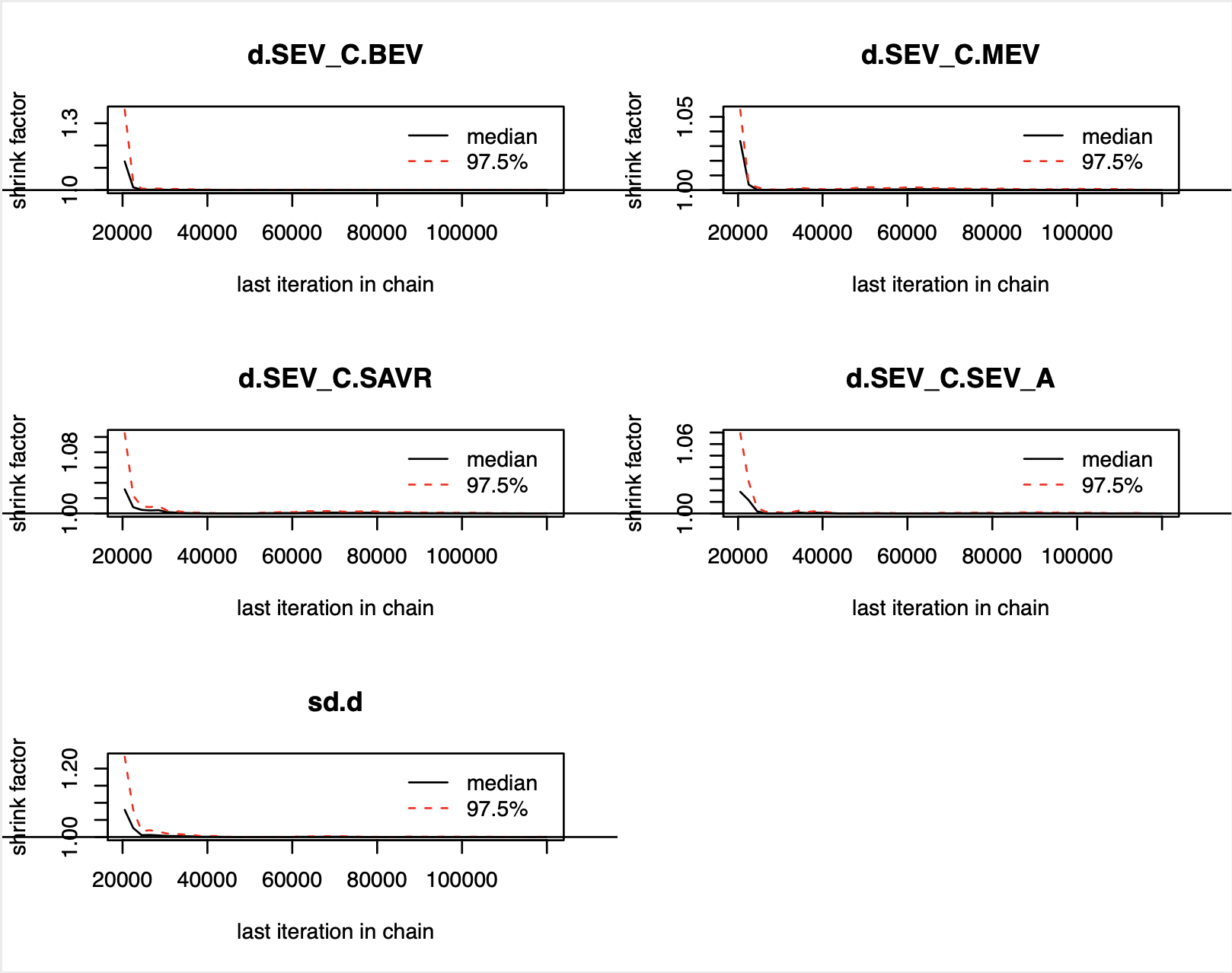

Figure 8S – Gelman-brooks-rubin convergence plot for stroke at 30 days. Abbreviations same as figure 1S.

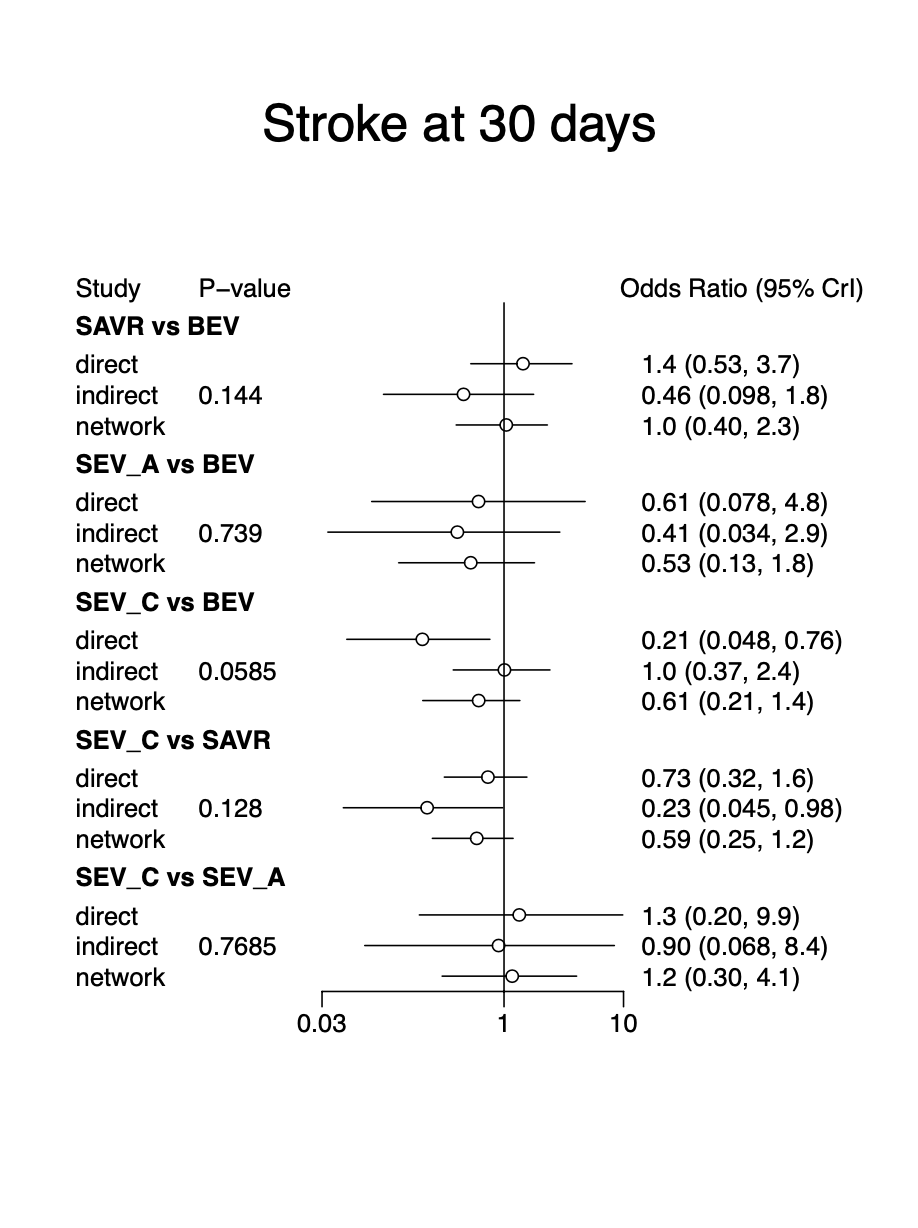

Figure 9S– Node splitting model for stroke at 30 days after various TAVR valves. Abbreviations same as figure 1S.

### Convergence diagnostics for stroke at 1 year

Potential scale reduction factors:

Point est. Upper C.I.

d.SEV_C.BEV	1	1

d.SEV_C.MEV	1	1

d.SEV_C.SAVR	1	1

d.SEV_C.SEV_A	1	1

sd.d	1	1

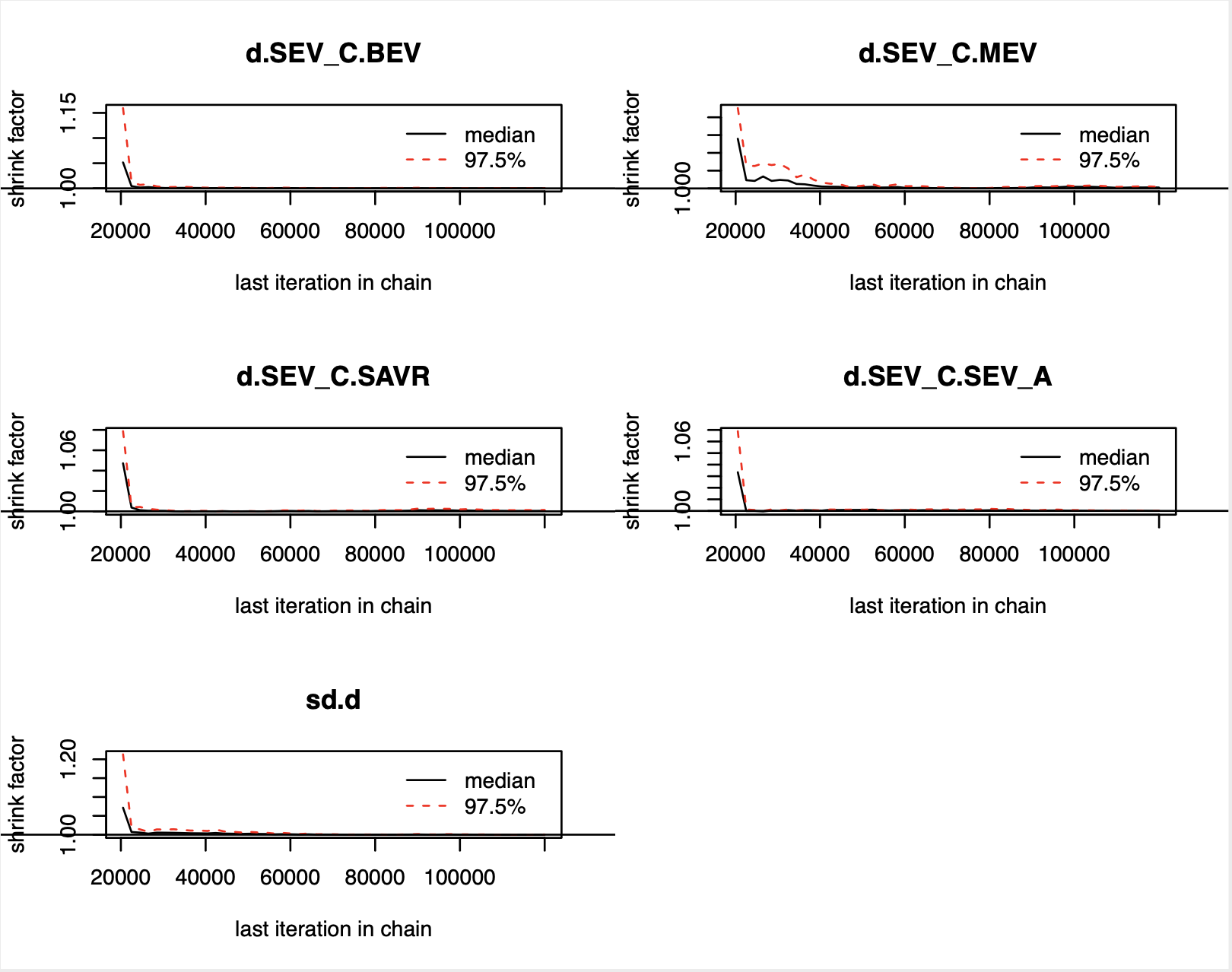

Figure 10S – Gelman-brooks-rubin convergence plot for stroke at one year. Abbreviations same as figure 1S.

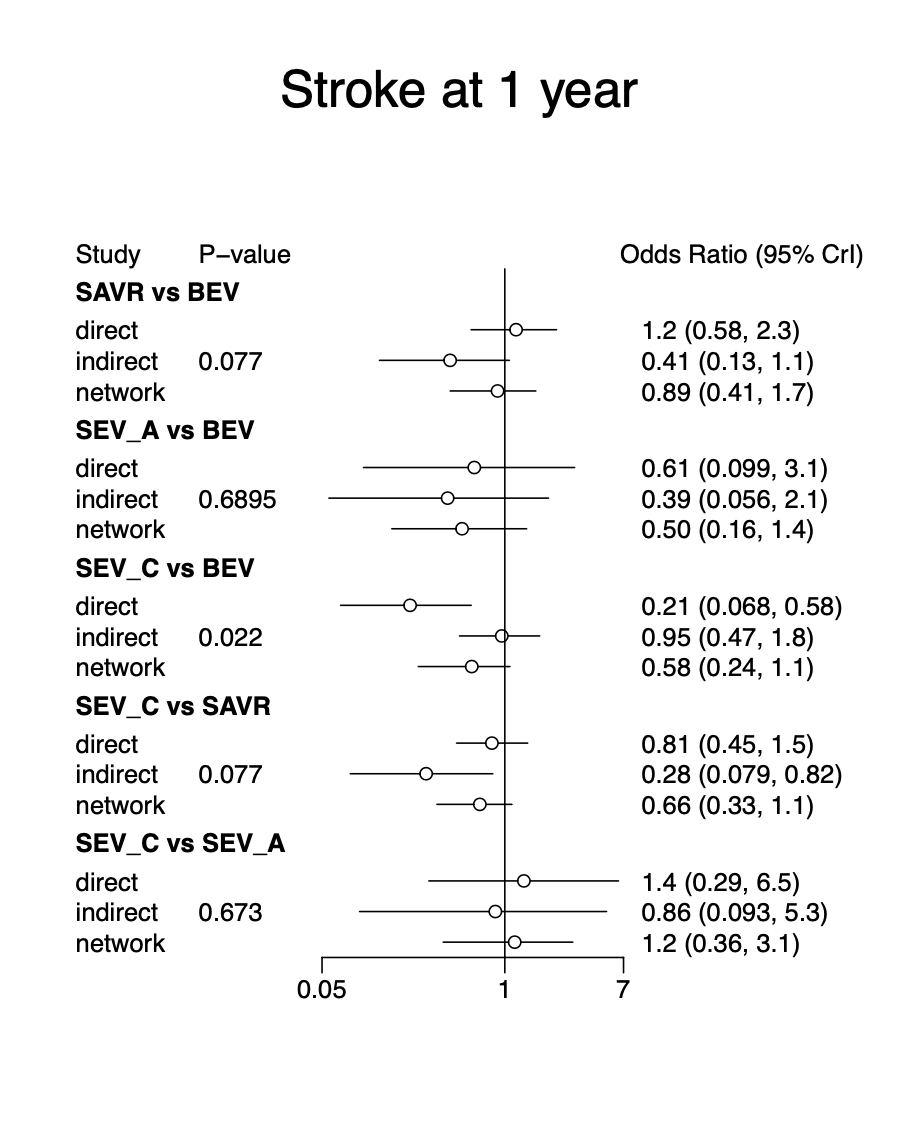

Figure 11S– Node splitting model for stroke at one year after various TAVR valves. Abbreviations same as figure 1S.

### Convergence diagnostic for Moderate or more aortic regurgitation at 30 days

Potential scale reduction factors:

Point est. Upper C.I.

d.SEV_C.BEV	1	1

d.SEV_C.MEV	1	1

d.SEV_C.SAVR	1	1

d.SEV_C.SEV_A	1	1

sd.d	1	1

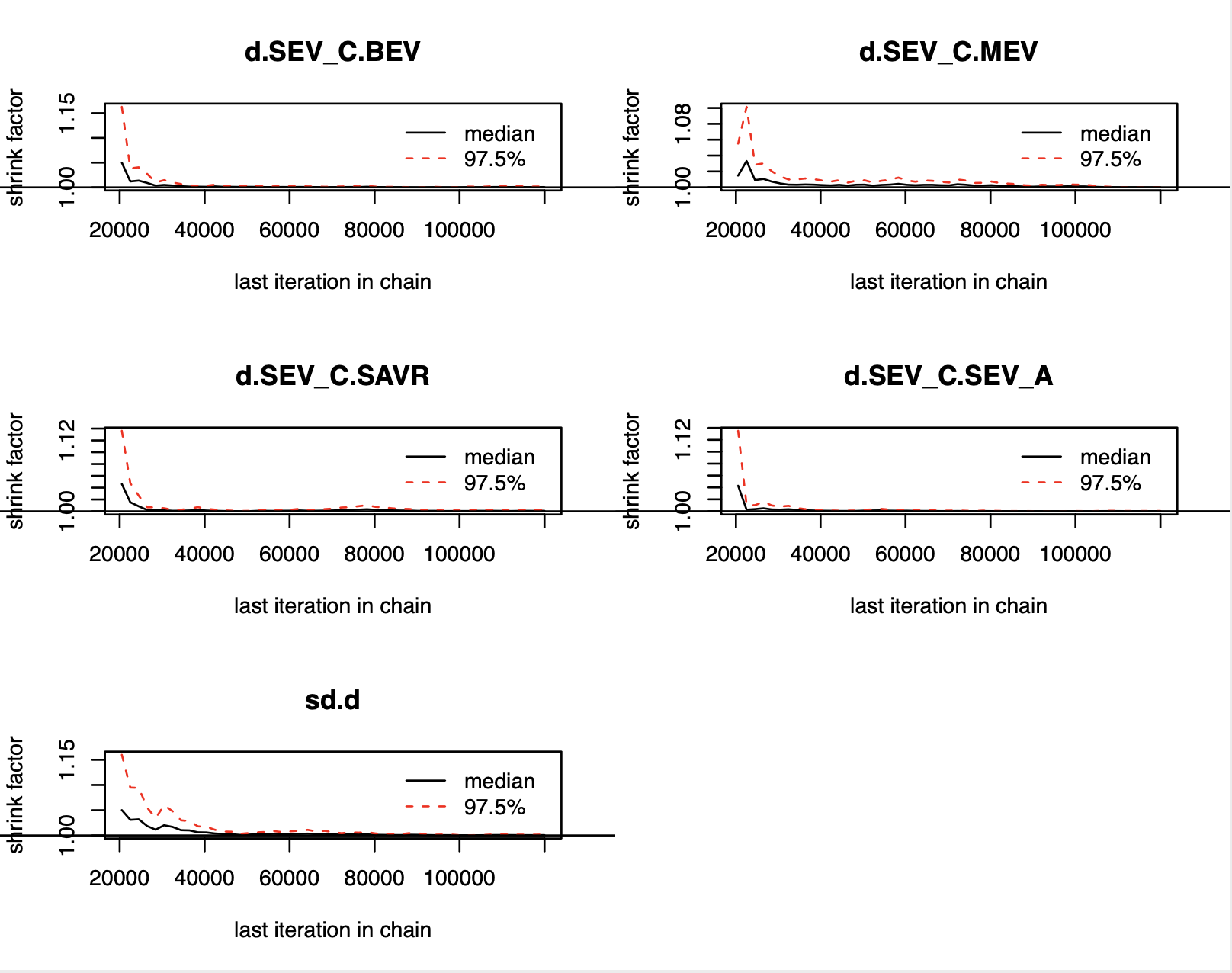

Figure 12S – Gelman-brooks-rubin convergence plot for moderate or more aortic regurgitation at 30 days. Abbreviations same as figure 1S.

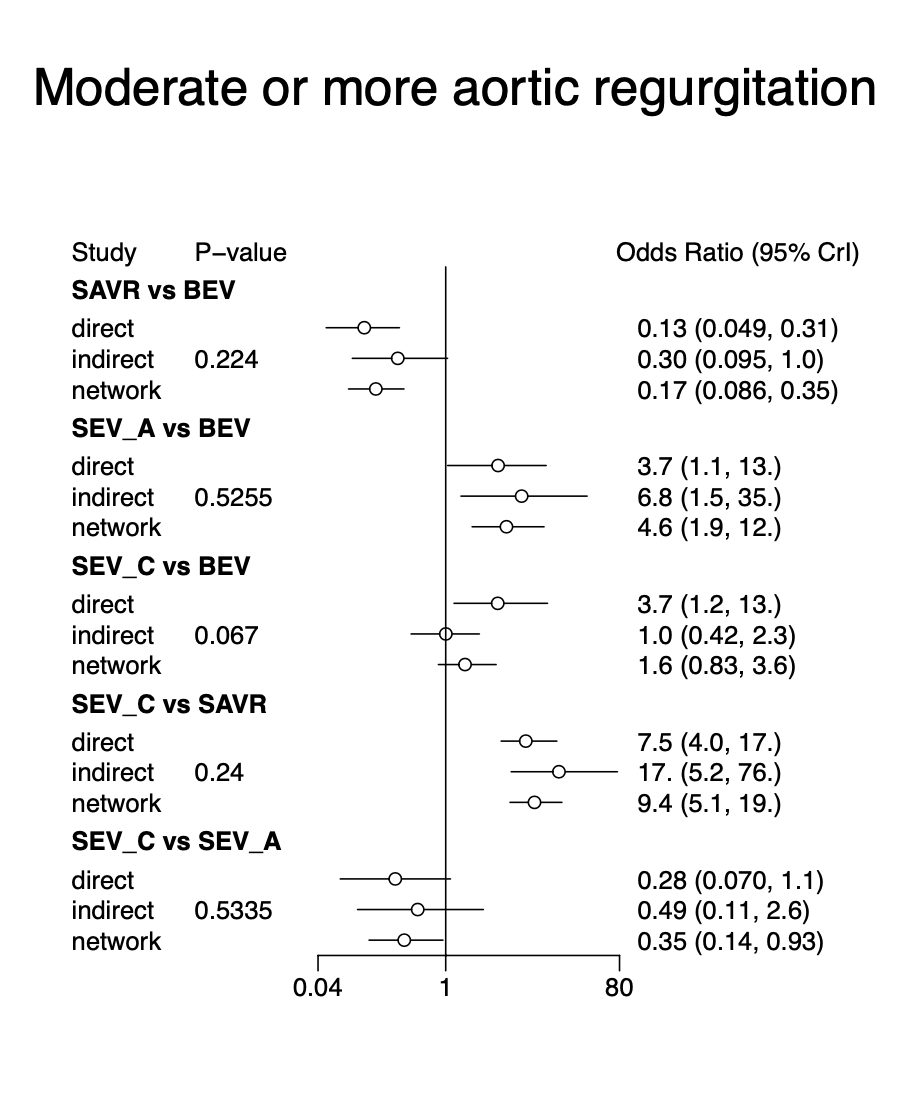

Figure 13S - Node splitting model for moderate or more aortic regurgitation at 30 days after various valve interventions. Abbreviations same as figure 1S.

### Convergence diagnostics for moderate or more aortic regurgitation at 1 year

Potential scale reduction factors:

Point est. Upper C.I.

d.SEV_C.BEV	1	1

d.SEV_C.MEV	1	1

d.SEV_C.SAVR	1	1

d.SEV_C.SEV_A	1	1

sd.d	1	1

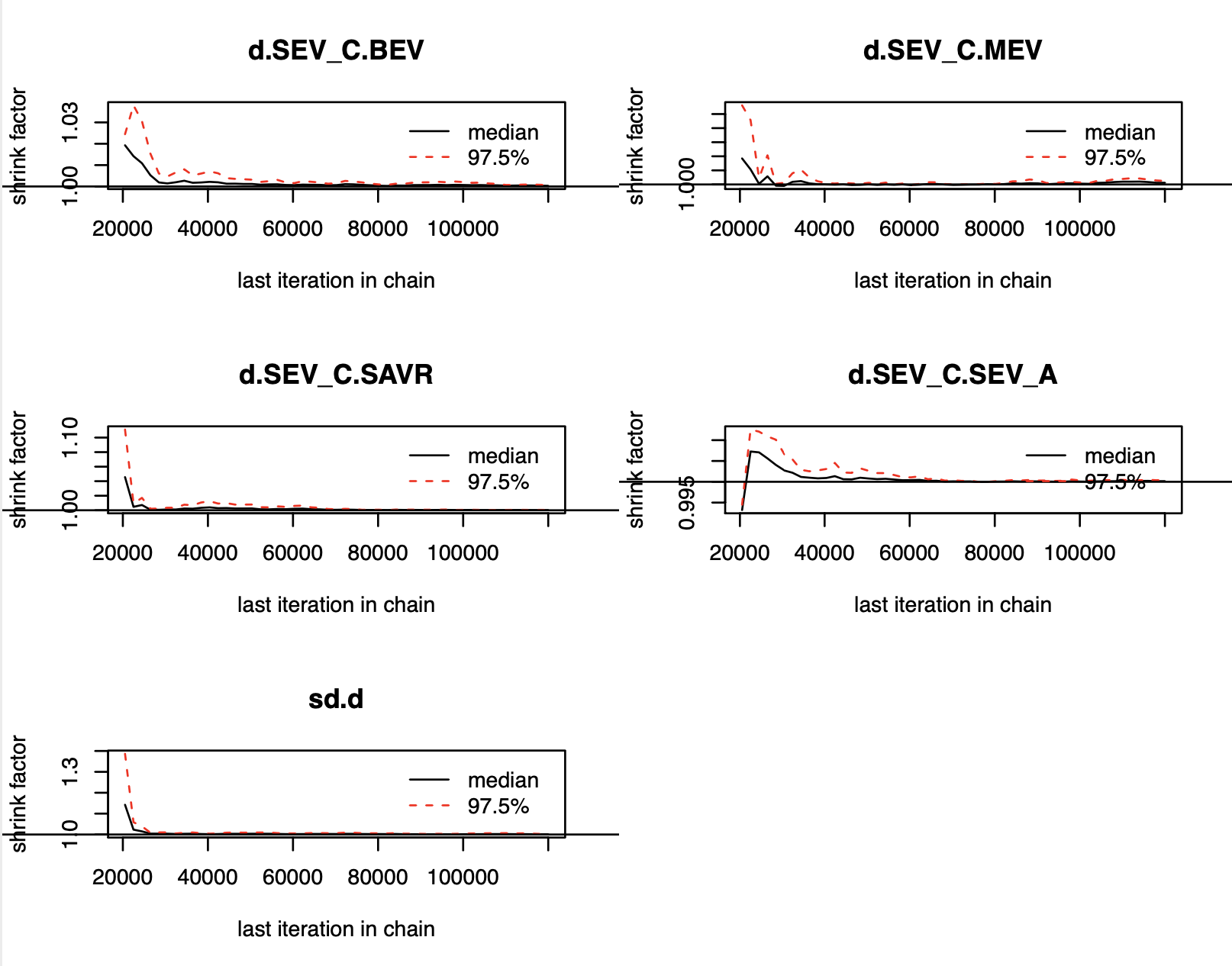

Figure 14S – Gelman-brooks-rubin convergence plot for moderate or more aortic regurgitation at 1 year. Abbreviations same as figure 1S.

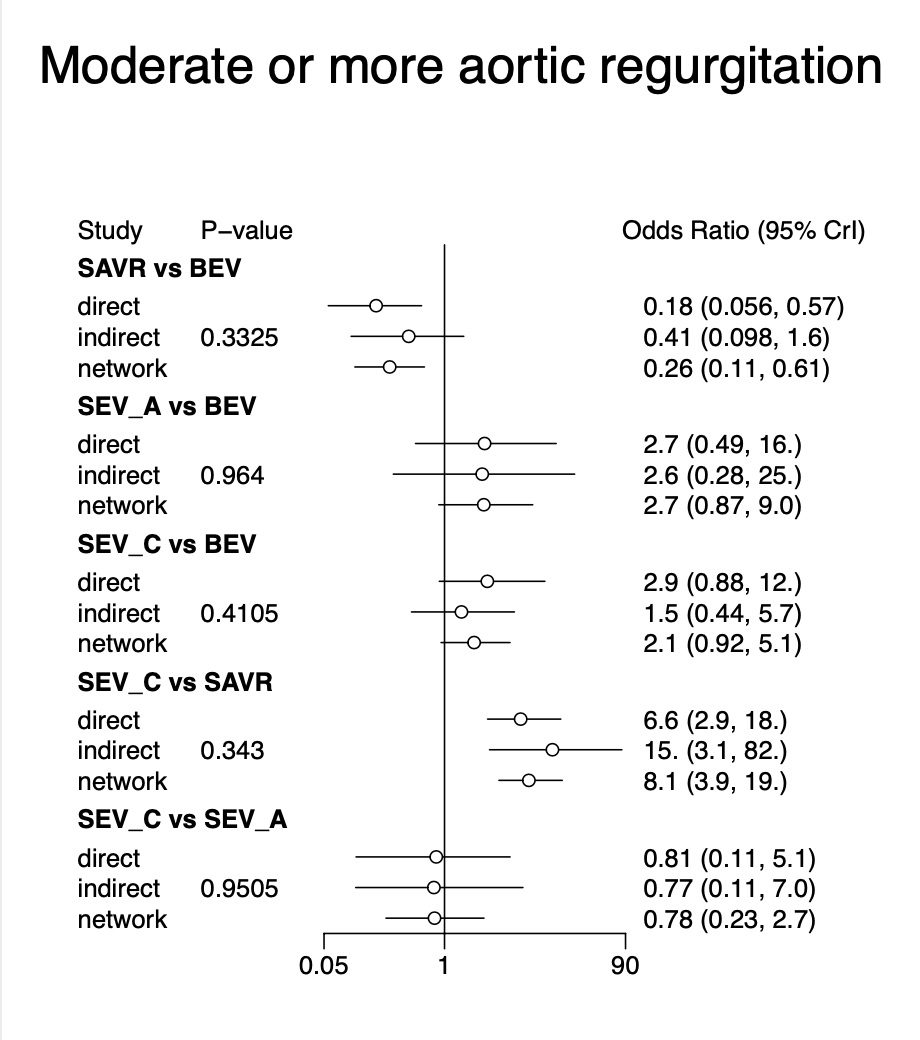

Figure 15S - Node splitting model for moderate or more aortic regurgitation at 1 year after various valve interventions. Abbreviations same as figure 1S.

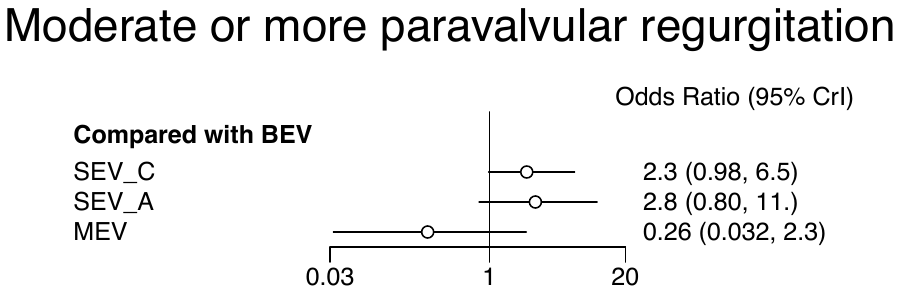

Figure 16S – Forest plot showing odds of moderate or more aortic regurgitation at 1 year after SEV_C, SEV_A and MEV compared BEV. Abbreviations same as Figure 1S.

### Convergence diagnostic for pacemaker implantation at 30 days

Potential scale reduction factors:

Point est. Upper C.I.

d.SEV_C.BEV	1	1

d.SEV_C.MEV	1	1

d.SEV_C.SAVR	1	1

d.SEV_C.SEV_A	1	1

sd.d	1	1

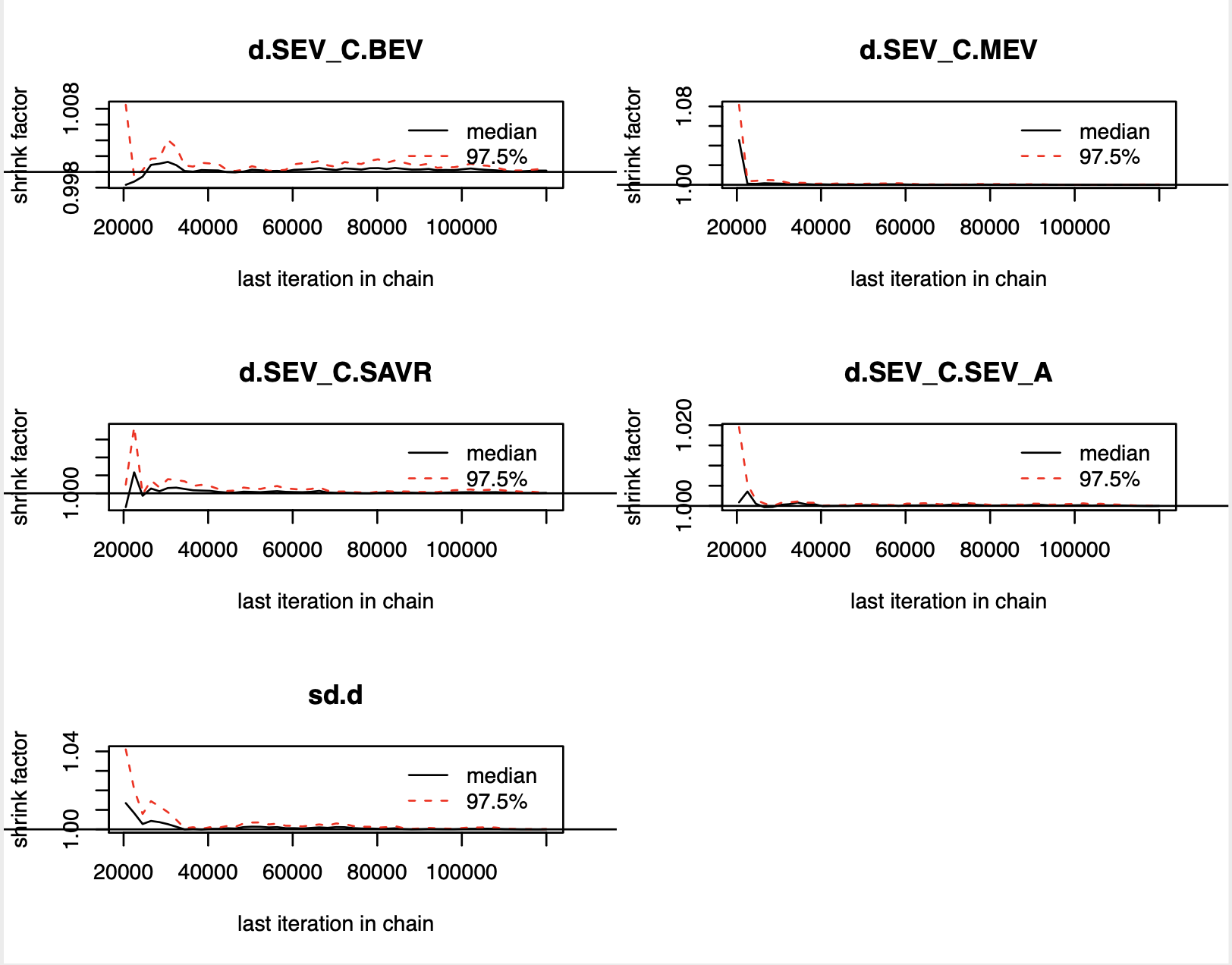

Figure 17S – Gelman-brooks-rubin convergence plot for pacemaker implantation at 30 days. Abbreviations same as figure 1S.

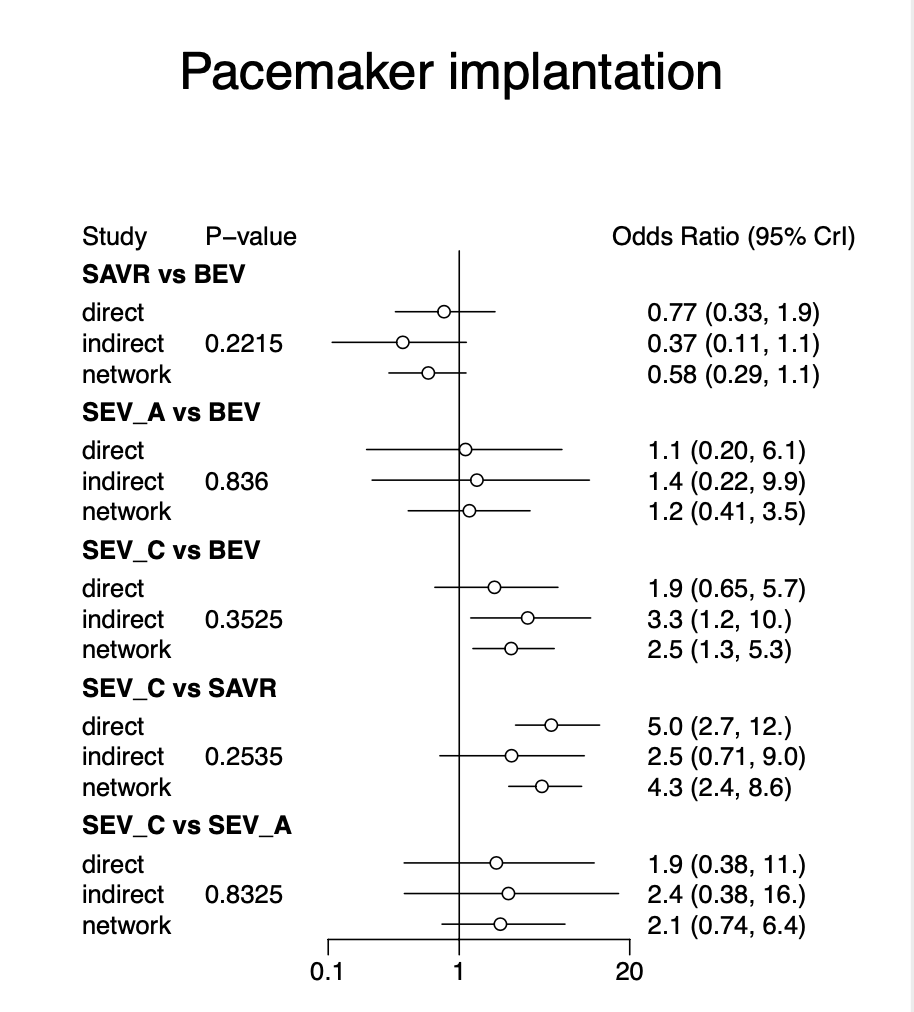

Figure18S - Node splitting model for pacemaker implantation at 30 days after various valve interventions. Abbreviations same as figure 1S.

### Gelman diagnostic for pacemaker implantation at 1 year

Potential scale reduction factors:

Point est. Upper C.I.

d.SEV_C.BEV	1	1

d.SEV_C.MEV	1	1

d.SEV_C.SAVR	1	1

d.SEV_C.SEV_A	1	1

sd.d	1	1

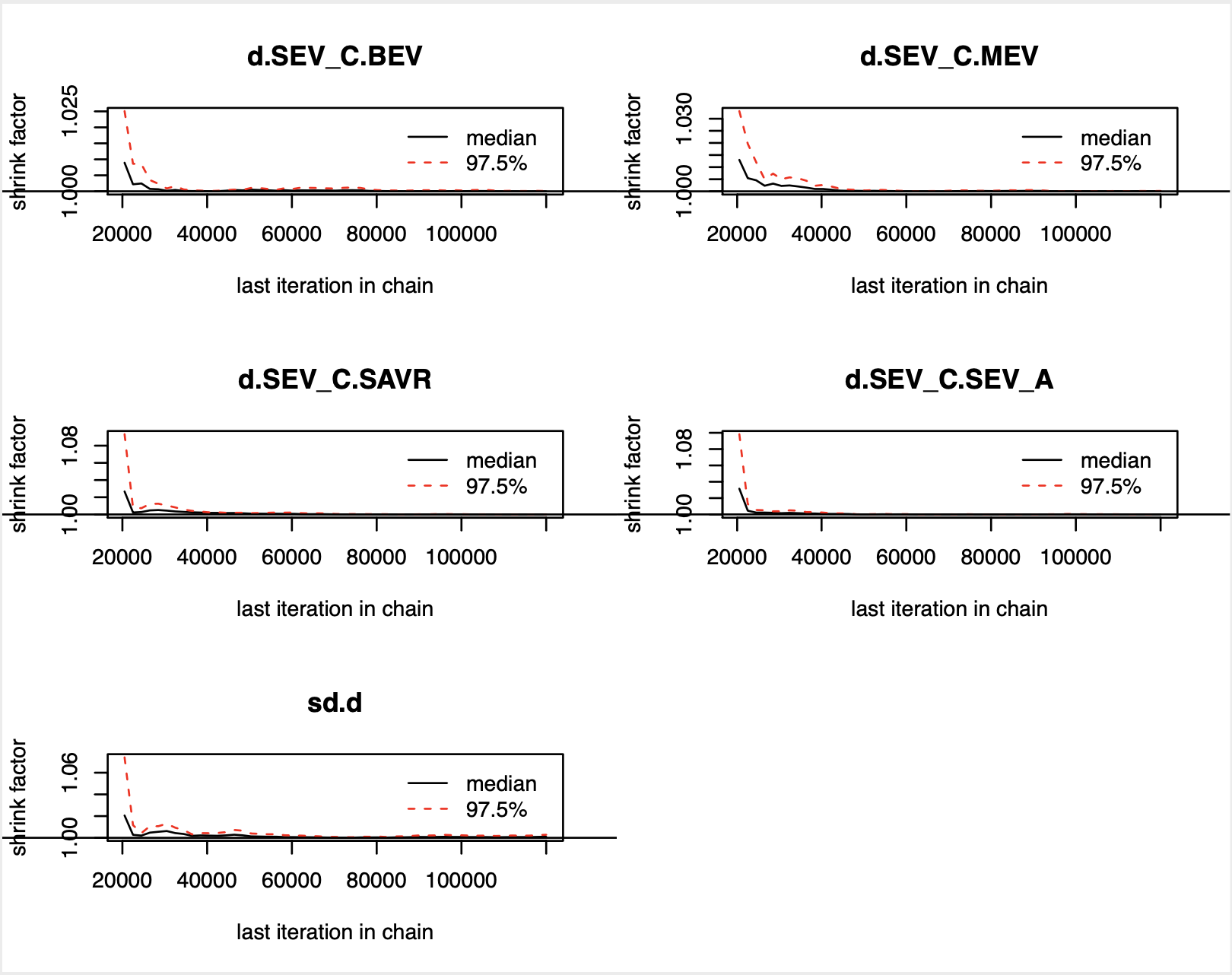

Figure 19S – Gelman-brooks-rubin convergence plot for pacemaker implantation at 1 year. Abbreviations same as figure 1S.

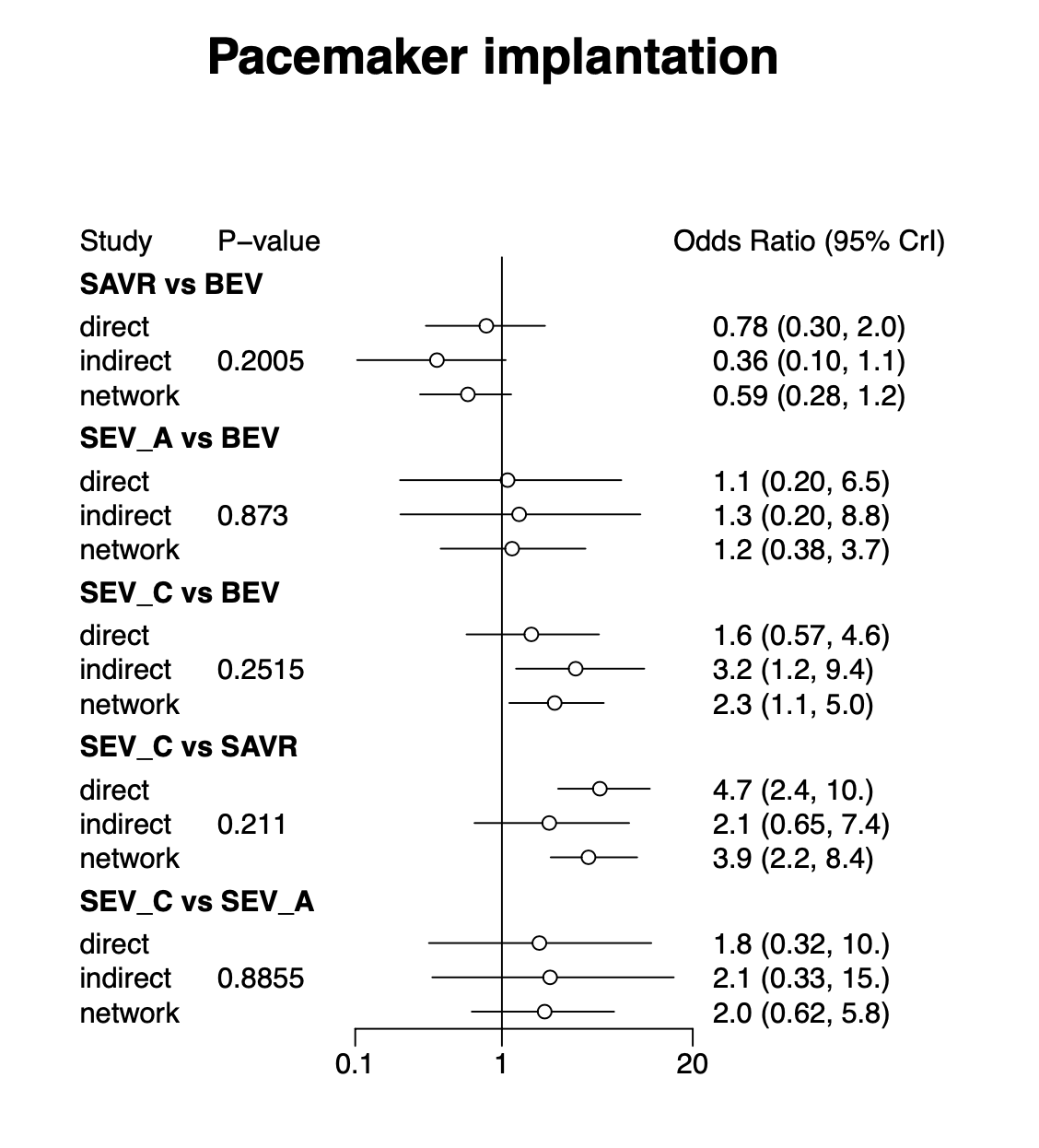

Figure20S - Node splitting model for pacemaker implantation at 1 year after various valve interventions. Abbreviations same as figure 1S.

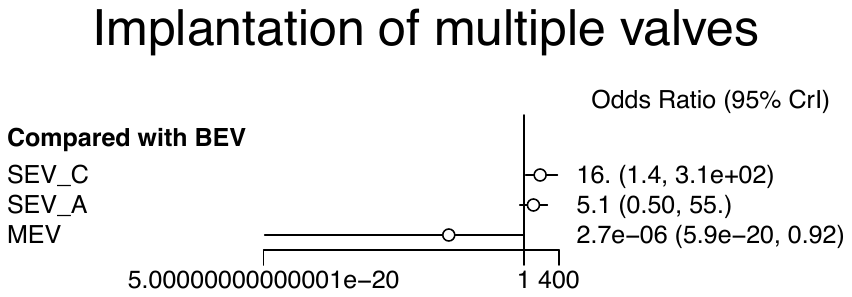

Figure 21 S - Forest plot showing comparison different TAVR valves for use > 1 valve during the procedure.

### Convergence diagnostics for multiple valve implantation

Potential scale reduction factors:

Point est. Upper C.I.

d.SEV_C.BEV	1.00	1.00

d.SEV_C.MEV	1.17	1.46

d.SEV_C.SEV_A	1.00	1.00

sd.d	1.00	1.00

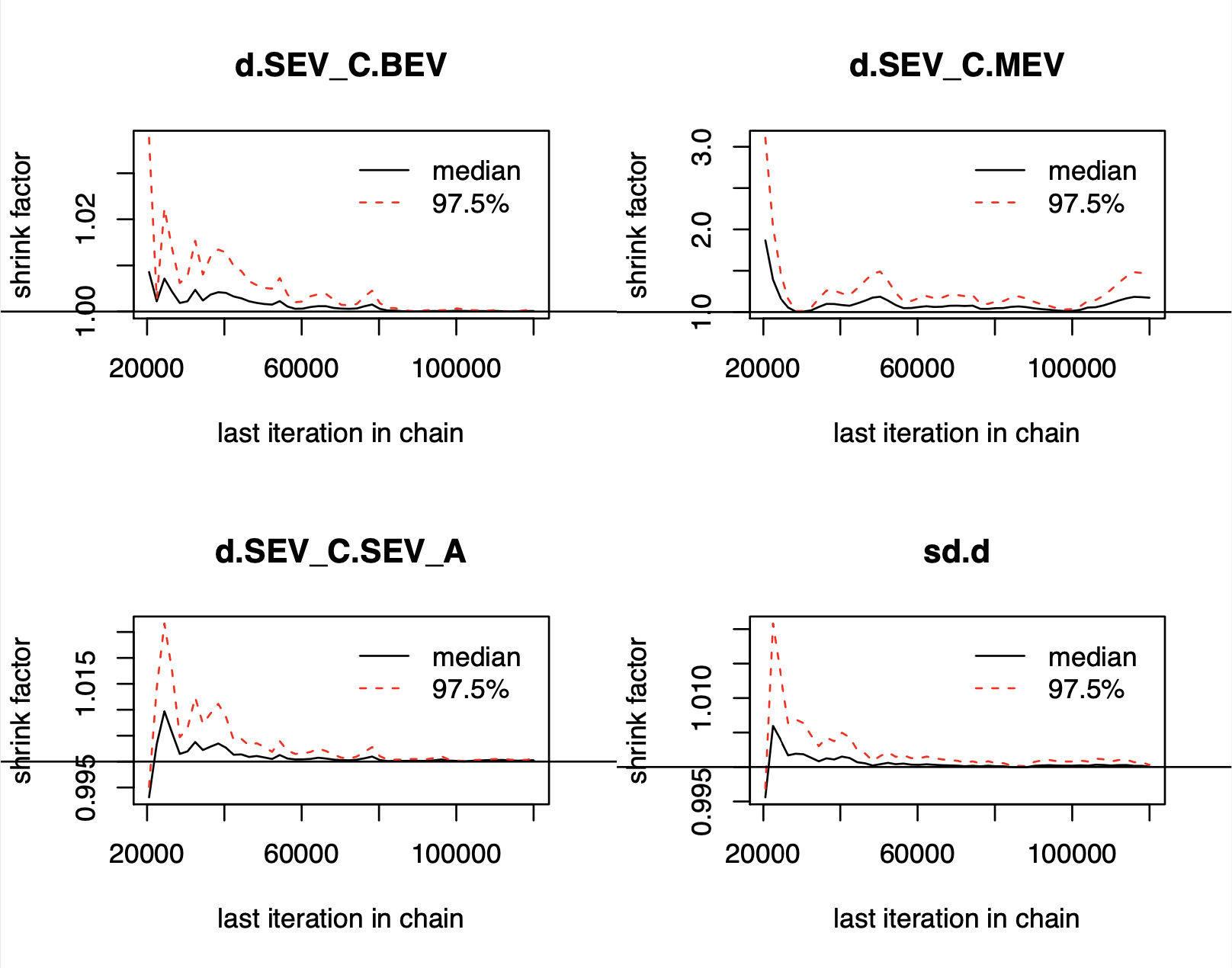

Figure 22S – Gelman-brooks-rubin convergence plot for > 1 TAVR valve implantation. Abbreviations same as figure 1S.

### Convergence Diagnostics for multiple valve implantation after excluding MEV trial

Potential scale reduction factors:

Point est. Upper C.I.

d.BEV.SEV_A	1	1

d.BEV.SEV_C	1	1

sd.d	1	1

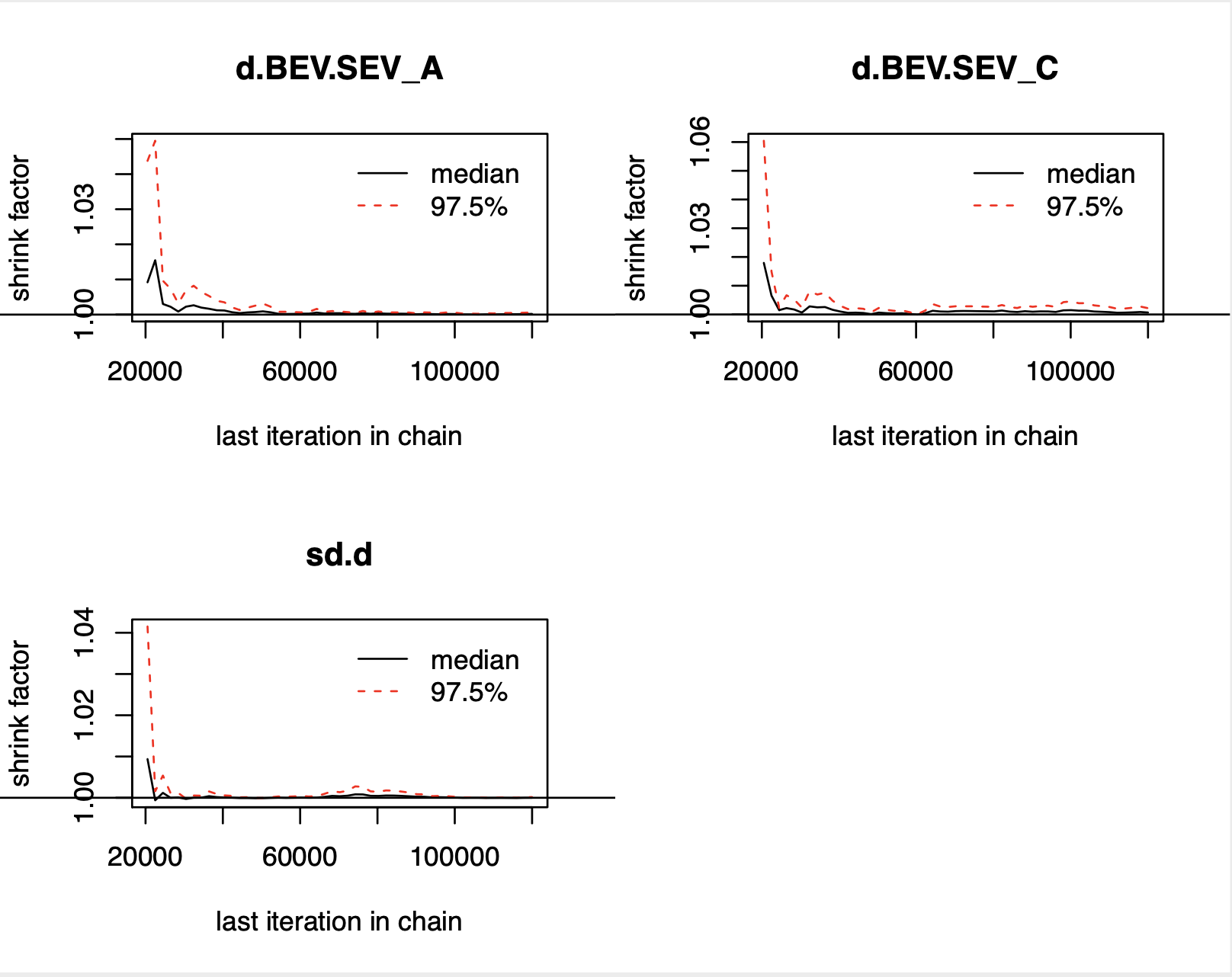

Figure 23S – Gelman-brooks-rubin convergence plot for > 1 TAVR valve implantation after excluding MEV trial. Abbreviations same as figure 1S.

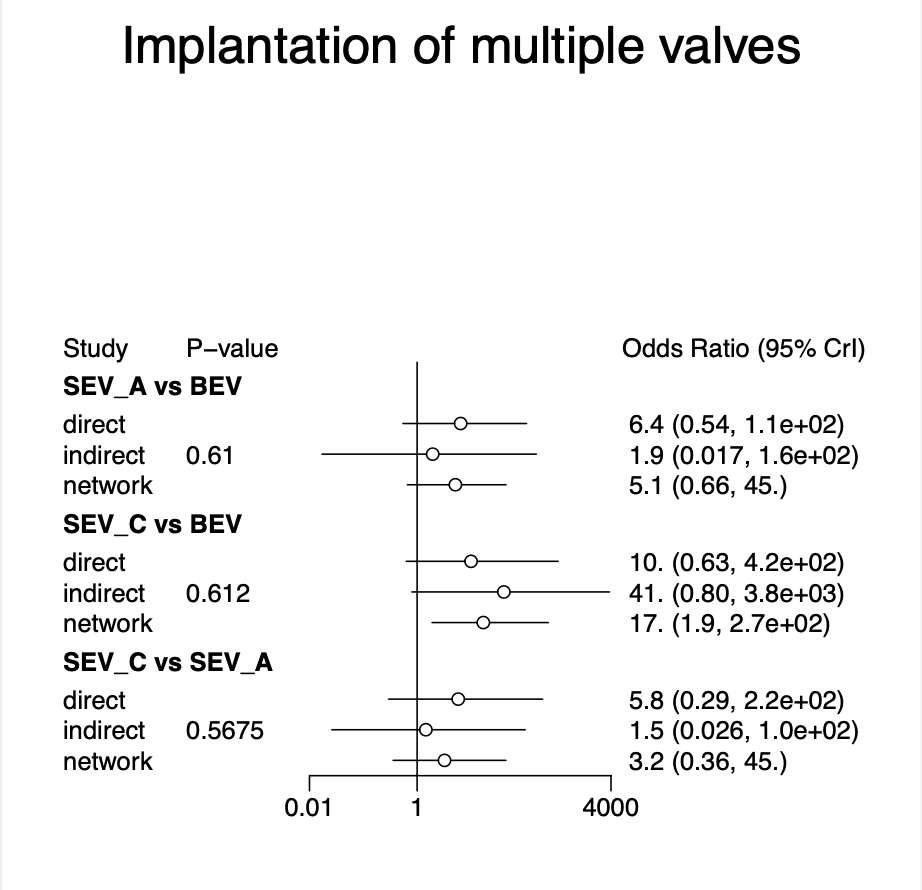

Figure 24S - Node splitting model for implantation multiple valves after various valve interventions.
