## Supplementary material for "Comparison of various transcatheter aortic valves for aortic stenosis – a network meta-analysis of randomized controlled trials": Table 1S

| **Study ID** | **Randomization process** | **Comment for randomization process** | **Deviations from intended interventions** | **Comment for deviations from intended interventions** | **Missing outcome data** | **Comment for missing outcome data** | **Measurement of the outcome** | **Comment for measurement of the outcome** | **Selection of the reported result** | **Comment for selection of the reported result** | **Overall Bias** | **Comment for overall bias** |
| --- | --- | --- | --- | --- | --- | --- | --- | --- | --- | --- | --- | --- |
| SCOPE I(5) | LOW | NONE | LOW | NONE | LOW | NONE | LOW | NONE | LOW | NONE | LOW | NONE |
| CHOICE(6) | LOW | NONE | LOW | NONE | LOW | NONE | LOW | NONE | LOW | NONE | LOW | NONE |
| SOLVE TAVI(7) | LOW | NONE | LOW | NONE | SOME CONCERNS | clinical follow up | LOW | NONE | LOW | NONE | LOW | NONE |
| SCOPE II(8) | LOW | NONE | LOW | NONE | SOME CONCERNS | echo follow up only | LOW | NONE | LOW | NONE | LOW | NONE |
| REPRISE III (9) | LOW | NONE | LOW | NONE | SOME CONCERNS | clinical | LOW | NONE | LOW | NONE | LOW | NONE |
| Evolut low risk trial (12) | LOW | NONE | LOW | NONE | LOW | NONE | LOW | NONE | LOW | NONE | LOW | NONE |
| NOTION (13) | LOW | NONE | LOW | NONE | LOW | NONE | LOW | NONE | LOW | NONE | LOW | NONE |
| SURTAVI (14) | LOW | NONE | LOW | NONE | LOW | NONE | LOW | NONE | LOW | NONE | LOW | NONE |
| PARTNER 3 (3) | LOW | NONE | LOW | NONE | LOW | NONE | LOW | NONE | LOW | NONE | LOW | NONE |
| PARTNER 2 (TF cohort) (2) | LOW | NONE | LOW | NONE | LOW | NONE | LOW | NONE | LOW | NONE | LOW | NONE |
| PARTNER 1A (TF cohort) (1) | LOW | NONE | LOW | NONE | LOW | NONE | LOW | NONE | LOW | NONE | LOW | NONE |
| US CoreValve high risk trial(15) | LOW | NONE | LOW | NONE | LOW | NONE | LOW | NONE | LOW | NONE | LOW | NONE |
