## Supplementary material for "Comparison of various transcatheter aortic valves for aortic stenosis – a network meta-analysis of randomized controlled trials": Table 2S

| Outcomes | Global I^2^ statistics |
| --- | --- |
| Mortality at 30 days | 6% |
| Mortality at 1 year | 5% |
| Stroke at 30 days | 7% |
| Stroke at 1 year | 6% |
| ≥Moderate AR at 30 days | 7% |
| ≥Moderate AR at 1 year | 7% |
| Pacemaker implantation at 30 days | 5% |
| Pacemaker implantation at 1 year | 5% |
| Use of >1 TAVR valve | 22% |
