## Supplementary material for "Comparison of various transcatheter aortic valves for aortic stenosis – a network meta-analysis of randomized controlled trials": Table 3S

**Which valve most suitable for Transcatheter Aortic Valve Replacement**

**Patient or population:** patients with native aortic stenosis
**Settings:** randomized controlled trials for elective procedure
**Interventions:** SEV_C(Self-expanding valve – Corevalve), SEV_A (Self-expanding valve – Accurate), and MEV (Mechanically expandable valve)
**Comparison:** BAV (Balloon expanding valve)

| **Outcomes** | **Effects and confidence in the effects.** | | |
| --- | --- | --- | --- |
|  | **SEV_C** | **SEV_A** | **MEV** |
| **Death at 30 days** | **OR 1.3** | **OR 3** | **OR 1.5** |
|  | (0.7 to 2.4) | (1.1 to 8.7) | (0.39 to 5.7) |
|  | **⊕⊕⊕⊕** **High** | **⊕⊕🌕🌕 Low** | **⊕⊕🌕🌕 Low** |
|  | confidence in estimate | confidence in estimate | confidence in estimate |
|  | based on context of trivial or no difference | based on imprecision | based on imprecision |
| **Death at 1 year** | **OR 1.0** | **OR 1.4** | **OR 0.87** |
|  | (0.73 to 1.3) | (0.92 to 2.2) | (0.47 to 1.6) |
|  | **⊕⊕⊕⊕ High** | **⊕⊕⊕🌕 Moderate** | **⊕⊕⊕🌕 Moderate** |
|  | confidence in estimate | confidence in estimate | confidence in estimate |
|  | based on context of trivial or no difference | based on imprecision | based on imprecision |
| **Stroke at 30 days**  | **OR 0.61** | **OR 0.52** | **OR 0.70** |
|  | (0.20 to 1.3) | (0.12 to 1.8) | (0.084 to 4.1) |
|  | **⊕⊕🌕🌕 Low** | **⊕⊕🌕🌕 Low** | **⊕⊕🌕🌕 Low** |
|  | confidence in estimate | confidence in estimate | confidence in estimate |
|  | based on incoherence and imprecision | based on imprecision | based on imprecision |
| **Stroke at 1 year**  | **OR 0.58** | **OR 0.50** | **OR 0.43** |
|  | (0.24 to 1.1) | (0.15 to 1.4) | (0.078 to 1.7) |
|  | **⊕⊕🌕🌕 Low** | **⊕⊕🌕🌕 Low** | **⊕⊕🌕🌕 Low** |
|  | confidence in estimate | confidence in estimate | confidence in estimate |
|  | based on incoherence and imprecision | based on imprecision | based on imprecision |
| **≥ Moderate aortic regurgitation at 30 days**  | **OR 1.6** | **OR 4.6** | **OR 0.10** |
|  | (0.81 to 3.5) | (2 to 12) | (0.016 to 0.53) |
|  | **⊕⊕🌕🌕 Low** | **⊕⊕⊕🌕 Moderate** | **⊕⊕⊕🌕 Moderate** |
|  | confidence in estimate | confidence in estimate | confidence in estimate |
|  | based on risk of bias, incoherence, and imprecision | based on risk of bias and imprecision | based on imprecision |
| **≥ Moderate aortic regurgitation at 1 year**  | **OR 2.1** | **OR 2.7** | **OR 0.24** |
|  | (0.95 to 5.1) | (0.83 to 8.7) | (0.033 to 1.6) |
|  | **⊕⊕⊕🌕 Moderate** | **⊕⊕⊕🌕 Moderate** | **⊕⊕🌕🌕 Low** |
|  | confidence in estimate | confidence in estimate | confidence in estimate |
|  | based on risk of bias and imprecision | based on risk of bias and imprecision | based on imprecision |
| **Pacemaker implantation at 30 days**  | **OR 2.5** | **OR 1.2** | **OR 5.7** |
|  | (1.3 to 5.3) | (0.43 to 3.6) | (1.3 to 29) |
|  | **⊕⊕⊕⊕ High** | **⊕⊕⊕🌕 Moderate** | **⊕⊕⊕🌕 Moderate** |
|  | confidence in estimate | confidence in estimate | confidence in estimate |
|  | based on coherence and crossing of threshold | based on imprecision | based on imprecision |
| **Pacemaker implantation at 1 year**  | **OR 2.3** | **OR 1.2** | **OR 5.4** |
|  | (1.1 to 4.9) | (0.39 to 3.6) | (1.1 to 28) |
|  | **⊕⊕⊕⊕ High** | **⊕⊕⊕🌕 Moderate** | **⊕⊕⊕🌕 Moderate** |
|  | confidence in estimate | confidence in estimate | confidence in estimate |
|  | based on coherence and crossing of threshold | based on imprecision | based on imprecision |
| **Use > 1 TAVR valve**  | **OR 17** | **OR 5.1** | **NA** |
|  | (1.5 to 3e2) | (0.49 to 56) |  |
|  | **⊕⊕⊕🌕 Moderate** | **⊕⊕🌕🌕 Low** |  |
|  | confidence in estimate | confidence in estimate |  |
|  | based on imprecision but crossing of threshold | based on imprecision |  |

CI: Confidence interval; OR: Odds ratio;

GRADE Working Group grades of evidence
**High quality:** Further research is very unlikely to change our confidence in the estimate of effect.
**Moderate quality:** Further research is likely to have an important impact on our confidence in the estimate of effect and may change the estimate.
**Low quality:** Further research is very likely to have an important impact on our confidence in the estimate of effect and is likely to change the estimate.
**Very low quality:** We are very uncertain about the estimate.
